# AToP: An Explainability Framework for Transformer-Based EHR Predictions Using Attribution-Guided Temporal Pattern Mining

**DOI:** 10.64898/2026.11.10.26367753

**Authors:** Anas Belouali, Hadi Kharrazi

## Abstract

**Objective:** To develop and evaluate AToP (Attribution to Temporal Patterns), a post-hoc explainability framework that converts per-token Integrated Gradients (IG) from a Transformer into structured temporal pattern narratives.

**Materials and Methods:** AToP was applied to a single-stream Transformer trained on MIMIC-IV v3.1 (218,196 patients; 12.6% unplanned 30-day readmission). It mines frequent episodes from salient-token subsequences in the training cohort, aggregates attribution among patients whose records contain each token or pattern, and collapses near-duplicates by patient overlap. Validation used masking, reversed-order analysis, token-level faithfulness tests, and KernelSHAP comparison.

**Results:** The Transformer achieved AUROC 0.77 and PR-AUC 0.34. Population-weighted attribution emphasized high-prevalence codes with small per-patient impact. Carrier-conditional attribution, the mean among patients with the token, surfaced rare high-attribution psychiatric markers including psychosis, self-harm overdose, suicidal ideation (SI), and severe major depressive disorder (MDD). Pattern-level attribution identified within-visit co-occurrences and cross-visit trajectories not captured by token rankings. Risk-increasing patterns included psychiatric sequences ({MDD, SI} → SI) and oncologic progressions (cancer anemia → ascites). Obstetric and chest pain evaluation patterns were associated with lower predicted readmission.

**Discussion:** The framework provides two complementary views of model predictions: carrier-conditional aggregation highlights rare, highly attributed clinical features, while temporal pattern analysis identifies within-visit combinations and cross-visit trajectories missed by token-level rankings. Masking and visit-order reversal provided evidence of correspondence with model behavior, but pattern attributions do not represent unique effects or establish clinical validity.

**Conclusion:** AToP turns a Transformer’s attributions into visit-anchored temporal pattern narratives, showing which events, in which order, contribute to its readmission predictions.

## Background and Significance

Transformer-based models have increasingly been used to learn directly from longitudinal electronic health record (EHR) data, from architectures such as BEHRT^1^ and Med-BERT^2^ to more recent longitudinal EHR foundation models.^3,4^ These approaches have been applied across diverse clinical prediction tasks.^5,6^ However, the same capability to model dependencies across heterogeneous clinical events creates an interpretability gap: how those events combine over time to produce a prediction remains difficult to discern.

As predictive models become more widely used in clinical care, clinicians, regulators, and institutional review processes increasingly emphasize explanations that are locally faithful, showing which inputs drove an individual prediction, and globally coherent, showing recurring patterns in model behavior across patients. Existing post-hoc attribution methods, including SHAP,^7^ IG,^8^ and DeepLIFT,^9^ assign importance scores to individual input features. In longitudinal EHR applications, these scores are often summarized across patients as rankings of clinical events.^10–12^ Such rankings do not capture which events co-occur within visits or how events are ordered across visits. Attention-based models such as RETAIN^13^ offer a partial alternative by reflecting temporal order through visit- and feature-level importance. However, the faithfulness of attention weights as explanations remains debated.^14,15^ Interaction attribution methods, such as SHAP interaction values^16^ and Integrated Hessians,^17^ quantify pairwise feature interactions but do not represent ordered sequences of events across visits.

The lack of temporal and combinatorial structure in conventional attribution summaries is especially consequential in longitudinal clinical data, where the same clinical events may carry different meaning depending on whether they occur together within an encounter or sequentially across encounters. To address this limitation, we propose using sequential pattern mining to organize model-attributed clinical events into recurring within-visit combinations and ordered cross-visit sequences. Prior work has applied sequential pattern mining to longitudinal EHR and claims data to study disease progression, adverse drug events, treatment pathways, and diagnostic trajectories.^18–23^ In related work combining temporal pattern mining and explainability, sequential pattern mining has been used to construct inputs for Transformer-based disease trajectory prediction, with SHAP subsequently applied to explain individual predictions.^24^ However, that approach uses mined patterns as predictive inputs rather than using model-derived attributions to guide temporal pattern discovery and explain model behavior.

We present AToP (Attribution to Temporal Patterns), a post-hoc explainability framework that combines token-level attribution from a trained Transformer with sequential pattern mining to probe how combinations and sequences of clinical events contribute to model predictions. We evaluate AToP on unplanned 30-day hospital readmission, a prediction task for which conventional risk models have historically shown modest discrimination.^25^

### Objective

To develop and evaluate AToP, a post-hoc explainability framework that uses token-level attribution to guide sequential pattern mining and examine how clinical events combine and unfold over time in Transformer-based EHR predictions.

## Materials and Methods

### Data and prediction task

The study cohort included 218,196 patients from MIMIC-IV v3.1,^26,27^ with an unplanned 30-day hospital readmission rate of 12.6%. The prediction target was unplanned readmission within 30 days of index discharge. The index admission was defined as the patient’s penultimate hospitalization. All earlier admissions were used as longitudinal history. For patients with a single admission, that admission served as the index. These 119,874 patients (54.9%) had no subsequent admission recorded in MIMIC-IV and were labeled as not readmitted. Among patients with two or more admissions, the readmission rate was 28.0%.

Each patient’s longitudinal record through the index admission was represented as a single chronological token sequence. The sequence contained ICD-9 and ICD-10 diagnosis and procedure codes, along with prescription drugs. ICD-9 codes were mapped to ICD-10 using the CMS General Equivalence Mappings,^28^ retaining only unambiguous one-to-one mappings. Codes without such a mapping were kept in their original ICD-9 form. Visit boundaries were marked with explicit [VISIT] tokens. Each admission formed one visit block, with blocks ordered chronologically. The sequence encodes the order of admissions, not the elapsed time between them. Within a block, tokens were grouped by type, and the pattern analysis treated events within a visit as unordered co-occurrences.

Chronic conditions were retained only at first appearance or when recorded as the primary diagnosis. Drugs were retained only at first recorded prescription, and ingredients prescribed in more than 25% of admissions were excluded to prevent routine inpatient treatments from dominating the mined patterns. Prescription drugs were mapped to RxNorm ingredient concepts using the RxNav API, by National Drug Code and then by drug name. Tokens occurring fewer than twice in the training set were excluded, giving a 24,005-token vocabulary of diagnoses, procedures, RxNorm ingredients, and the [VISIT], [CLS], and [PAD] tokens (**Supplementary Methods**).

Unplanned readmissions included emergency, urgent, direct, and observation admissions within 30 days of index discharge. Elective and surgical same-day admissions were excluded. Admissions ending in in-hospital death (11,801; 2.2%) were removed before index selection and labeling. Among index admissions, 709 (0.32%) were followed only by an admission ending in death. These were labeled as not readmitted. This yielded 27,508 unplanned 30-day readmissions.

### Single-stream Transformer

A Transformer encoder^29^ with a 24,005-token vocabulary and approximately 3.5 million parameters was trained using class-weighted binary cross-entropy loss. The architecture included two encoder layers, four attention heads, 128-dimensional embeddings, and 512-dimensional feed-forward layers. Early stopping was based on validation AUROC. All hyperparameters are listed in **Table S1**.

Diagnoses, procedures, medications, and the [VISIT], [CLS], and [PAD] special tokens were represented in a shared embedding space. Token embeddings were summed with learned absolute positional embeddings to encode sequence order. No separate visit-level or elapsed-time embeddings were used. The [CLS] representation was passed through a two-layer feed-forward head (128 to 64 to 1 units) with sigmoid activation to produce the predicted readmission probability.

Patients were split at the patient level into training (70%, n=152,737), validation (10%, n=21,819), and test (20%, n=43,640) cohorts. IG attribution and AToP analyses included 43,602 test patients. The remaining 38 yielded no salient-token set.

### Baselines and performance evaluation

We compared the Transformer with two baselines trained on seven structured predictors: the four components of the LACE readmission index^30^ (length of stay, emergent admission, Charlson comorbidity index, and emergency department visits in the preceding six months), entered as raw values rather than LACE points, plus age, sex, and number of prior admissions. Logistic regression used standardized predictors with L2 regularization. XGBoost used 200 trees, a maximum depth of 4, a learning rate of 0.05, and positive-class weighting for class imbalance. Both baselines used fixed hyperparameters and were evaluated on the test cohort.

Discrimination was summarized by the area under the receiver operating characteristic curve (AUROC) and the area under the precision-recall curve (PR-AUC). Differences between models were estimated with a paired bootstrap of test-cohort patients (1,000 resamples), reporting percentile 95% confidence intervals and two-sided p values with add-one correction, for which the smallest attainable value is 0.002.

### AToP explanation framework

AToP operates on the trained model in five steps (**Figure 1**): (1) compute token-level attribution using Integrated Gradients; (2) select salient tokens by cumulative share of each patient’s absolute attribution; (3) mine frequent episodes from the training cohort’s salient tokens: single-token events, within-visit co-occurrences, and cross-visit sequences; (4) aggregate attribution at the token and pattern levels over carriers; and (5) collapse near-redundant patterns by Jaccard similarity of their carrier sets.

**Figure 1.**
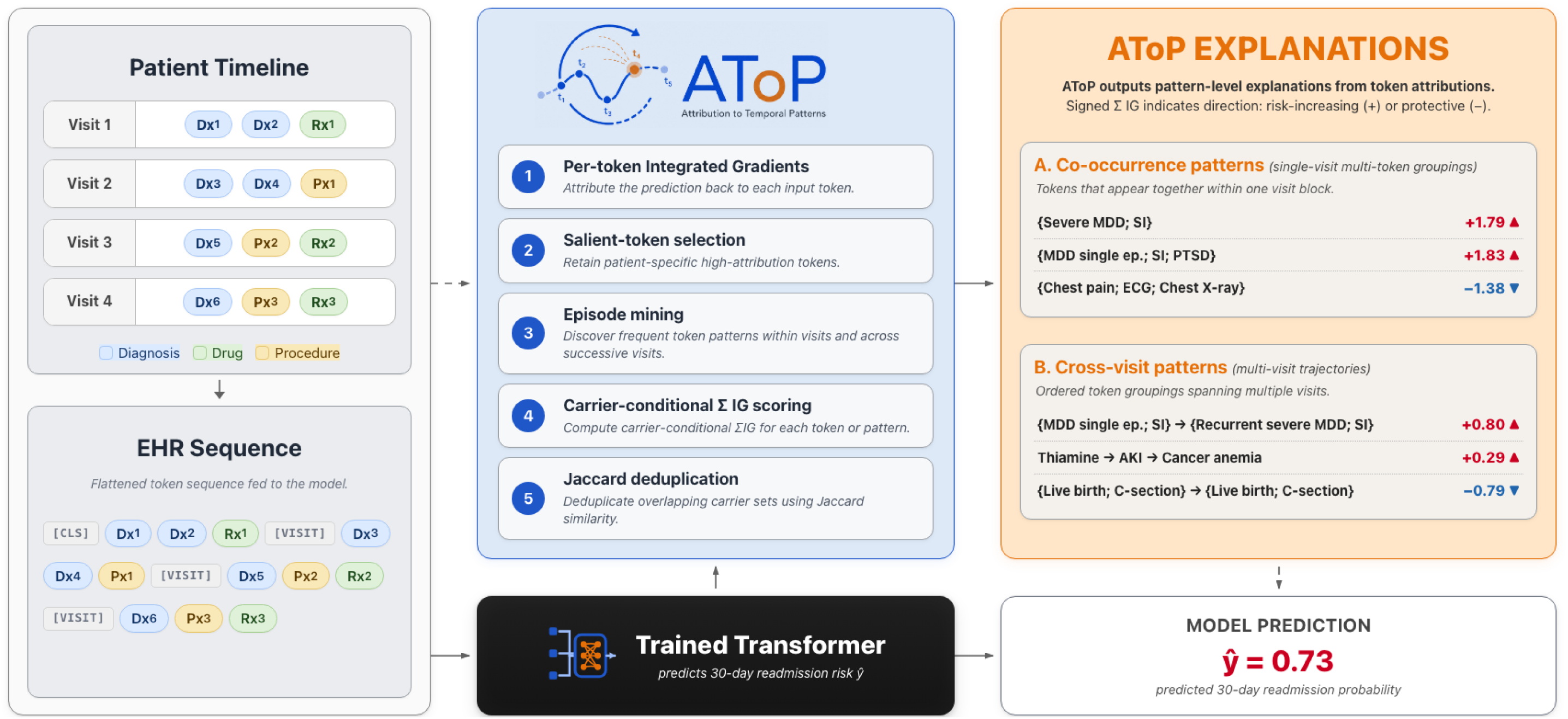
AToP framework overview. A trained Transformer predicts readmission from longitudinal EHR records. AToP identifies salient clinical tokens using Integrated Gradients (IG), mines recurring combinations within visits and sequences across visits, and summarizes their attribution among patients carrying each pattern. Jaccard similarity groups patterns with overlapping carrier sets. Predicted probabilities and attribution values are illustrative. MDD = major depressive disorder; SI = suicidal ideation; PTSD = post-traumatic stress disorder; ECG = electrocardiogram; AKI = acute kidney injury; ep. = episode.

We calculated Integrated Gradients on the model’s pre-sigmoid logit. By the completeness property, attributions over all input positions, including the [CLS] and [VISIT] tokens, sum approximately to the difference between the patient’s logit and that of an all-[PAD] reference sequence.^8,31^ Attribution signs are relative to this reference: positive values indicate a contribution toward a higher logit than the reference sequence, and negative (protective) values toward a lower one. AToP explanations and pattern analyses used only non-special-token attributions. Details of the reference sequence, special-token attribution, and approximation error are provided in the **Supplementary Methods**.

For each patient, a token’s attribution score was its signed IG value, and a multi-token pattern’s attribution score was the sum of the signed IG values of its constituent tokens. Rankings used absolute attribution magnitude, with the sign retained.

We define a carrier as a patient whose record contains the token or pattern being evaluated. The population-weighted attribution for token j was calculated as the mean signed IG across all patients:

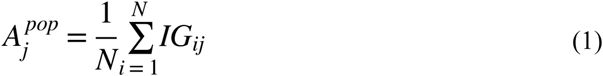

Carrier-conditional attribution was calculated as the mean signed IG among patients whose records contained token j:

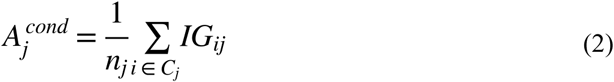

where *N* is the total number of patients, *C_j_* is the set of patients carrying token j, and *n_j_* is the number of carriers. Because noncarriers contributed zero attribution, the two measures are related by:

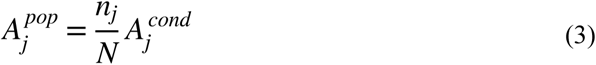

Tokens were ranked by the absolute magnitude of their respective attribution measures, with the sign retained to indicate direction. Equation (3) shows that population-weighted attribution reflects both token prevalence and attribution strength among carriers, whereas carrier-conditional attribution removes explicit prevalence weighting.

At the pattern level, patterns were matched step by step to each carrier’s full record, rather than only to the salient-token subsequence used for mining. Each step was assigned to the earliest visit containing all its tokens, with subsequent steps required to occur in strictly later visits. For each carrier, the pattern score was the sum of signed IG values for its tokens in the matched visits. These scores were averaged across carriers to obtain carrier-averaged ΣIG. For the per-patient display, each salient token occurrence was counted only once to avoid double-counting across overlapping patterns (**Supplementary Methods**).

Frequent-episode mining^18^ was implemented using a level-wise candidate-generation algorithm and applied only to salient-token subsequences from the training cohort. Patterns were constructed by either adding a co-occurring token within the same visit or appending a token from a later visit, yielding three pattern types: single-token events, within-visit co-occurrences, and cross-visit sequences. Mining was conducted separately in each outcome class. The resulting patterns were then pooled into a single set of distinct patterns that served as candidates for global rankings and per-patient explanations.

To reduce near-duplicate patterns, we applied greedy clustering based on Jaccard similarity between test-cohort carrier sets. Patterns were processed in descending order of absolute carrier-averaged ΣIG, with the highest-ranked pattern retained as the representative of each group.

Test-cohort carrier counts were used to determine reporting eligibility, and carrier-set overlap was used for deduplication. Neither influenced model training or pattern discovery.

### Parameter choices and sensitivity analyses

AToP parameter choices were guided by design and tractability constraints. We assessed sensitivity to several parameters, as described below and in the **Supplementary Methods**.

For mining, we retained for each patient the smallest set of tokens accounting for 80% of total absolute clinical-token IG, capped at 50 tokens. We assessed sensitivity to the attribution-mass threshold from 60% to 95%. Frequent-episode mining used a minimum support threshold of 0.1% of patients in each outcome class and a maximum of four tokens per pattern. Jaccard-based deduplication used a similarity threshold of 0.20, with sensitivity assessed across alternative thresholds.

After mining, separate eligibility criteria were applied to the all-pattern ranking, cross-visit ranking, and per-patient explanations. Cross-visit rankings were restricted to patterns spanning at least two visits and containing at least two distinct tokens. Minimum carrier counts for each ranking are provided in **Table S2**. Per-patient explanations used the subset of pooled patterns observed in at least 0.1% of patients in the full training cohort, a stricter floor than the class-specific mining threshold.

Additional implementation details are provided in the **Supplementary Methods**; parameter settings and their selection rationale are summarized in **Table S2**.

### Validation analyses

We evaluated AToP using pattern masking to assess effects on predicted readmission risk, reversed-order analysis to assess sensitivity to temporal order, token-level faithfulness tests to evaluate the influence of highly attributed tokens, and cross-method comparison with KernelSHAP to assess whether rankings depended on the attribution method.

For pattern masking, we selected the reported patterns and additional high-ranked patterns with positive or negative attribution, based on absolute carrier-averaged ΣIG. For each pattern, matched token positions were replaced with [PAD], and the change in predicted readmission probability was calculated as Δŷ = ŷ_original − ŷ_masked.

We assessed sensitivity to temporal order using two approaches. First, we compared pattern attribution between patients whose salient visit blocks contained a pattern in its original order and those whose blocks contained the reversed sequence. This analysis included the reported cross-visit patterns and additional patterns meeting minimum carrier-count criteria (**Supplementary Methods**). Second, we reversed the order of matched visits within patient records, recomputed IG, and compared pattern attribution before and after reversal.

We assessed token-level attribution faithfulness by measuring changes in predicted readmission probability after either masking the highest-ranked salient tokens or retaining those tokens while masking the remaining salient tokens.^32^

Comprehensiveness was measured as the absolute change in predicted readmission probability after masking the top k salient tokens ranked by absolute IG. For sufficiency, we retained the top k salient tokens, masked all remaining salient tokens, and measured the absolute change in predicted readmission probability. We varied k from 1 to 10, including only patients with at least k salient tokens. In both analyses, masking was restricted to salient clinical tokens, and non-salient and special tokens remained unchanged.

We compared both measures against a random baseline that selected the same number of salient tokens. Confidence intervals were estimated by bootstrapping patients. At k = 5, we compared comprehensiveness between IG-based and random token selection using a one-sided Wilcoxon signed-rank test.

To assess whether the rankings depended on the attribution method, we compared Integrated Gradients with KernelSHAP in the analytic test cohort. We compared population-weighted and carrier-conditional token rankings, as well as all-pattern and cross-visit pattern rankings. Pattern-level comparisons used carrier-conditional averaging (**Supplementary Methods**).

## Results

### Predictive performance

Cohort derivation is shown in **Figure S1**, and cohort characteristics are summarized in **Table 1**. The Transformer achieved an AUROC of 0.766 (95% CI, 0.759–0.773) and a PR-AUC of 0.342. Both LACE-based baselines had lower AUROC (XGBoost 0.728, logistic regression 0.659; both p = 0.002; **Table 2**).

**Table 1.** Cohort characteristics by unplanned 30-day readmission status.

| **Variable** | **Overall (N=218,196)** | **Readmitted (N=27,508, 12.6%)** | **Not readmitted (N=190,688, 87.4%)** | **SMD** |
|---|---|---|---|---|
| Age, median (IQR) | 57 (38–71) | 58 (40–72) | 57 (38–71) | 0.048 |
| Female sex, n (%) | 115,344 (52.9%) | 14,141 (51.4%) | 101,203 (53.1%) | 0.033 |
| Race/ethnicity, n (%) |  |  |  |  |
| White | 145,338 (66.6%) | 19,123 (69.5%) | 126,215 (66.2%) | 0.071 |
| Black | 28,373 (13.0%) | 3,836 (13.9%) | 24,537 (12.9%) | 0.032 |
| Hispanic or Latino | 11,922 (5.5%) | 1,360 (4.9%) | 10,562 (5.5%) | 0.027 |
| Asian | 9,543 (4.4%) | 1,344 (4.9%) | 8,199 (4.3%) | 0.028 |
| Other/Unknown | 23,020 (10.6%) | 1,845 (6.7%) | 21,175 (11.1%) | 0.155 |
| Admission type, n (%) |  |  |  |  |
| Emergency | 72,040 (33.0%) | 9,207 (33.5%) | 62,833 (33.0%) | 0.011 |
| Urgent | 25,351 (11.6%) | 2,684 (9.8%) | 22,667 (11.9%) | 0.069 |
| Observation | 98,109 (45.0%) | 12,961 (47.1%) | 85,148 (44.7%) | 0.049 |
| Elective | 22,696 (10.4%) | 2,656 (9.7%) | 20,040 (10.5%) | 0.028 |
| Length of stay (days), median (IQR) | 2.6 (1.0–5.1) | 3.2 (1.2–6.8) | 2.6 (1.0–4.9) | 0.178 |
| CCI, median (IQR) | 1 (0–2) | 1 (0–4) | 1 (0–2) | 0.36 |
| ED visits (prior 6 mo), n (%) |  |  |  |  |
| 0 | 129,156 (59.2%) | 16,451 (59.8%) | 112,705 (59.1%) | 0.014 |
| 1 | 74,138 (34.0%) | 7,063 (25.7%) | 67,075 (35.2%) | 0.208 |
| ≥2 | 14,902 (6.8%) | 3,994 (14.5%) | 10,908 (5.7%) | 0.295 |
| Number of admissions through index, n (%) |  |  |  |  |
| 1 | 163,271 (74.8%) | 12,316 (44.8%) | 150,955 (79.2%) | 0.758 |
| 2 | 19,800 (9.1%) | 4,899 (17.8%) | 14,901 (7.8%) | 0.302 |
| ≥3 | 35,125 (16.1%) | 10,293 (37.4%) | 24,832 (13.0%) | 0.585 |
| LACE score, median (IQR) | 7 (5–9) | 8 (5–11) | 7 (4–9) | 0.289 |

**Abbreviations:** CCI = Charlson Comorbidity Index; ED = emergency department; IQR = interquartile range; LACE = Length of stay, Acuity of admission, Charlson Comorbidity Index, and Emergency department visits; SMD = standardized mean difference. **Note:** CCI and admission counts include history through the index admission; prior ED visits cover the preceding six months.

**Table 2.** Predictive performance of the Transformer and baseline models.

| **Model** | **AUROC (95% CI)** | **PR-AUC (95% CI)** | **ΔAUROC (baseline − Transformer), 95% CI** | **P value** |
|---|---|---|---|---|
| Transformer | 0.766 (0.759, 0.773) | 0.342 (0.330, 0.355) | ref. |  |
| LACE XGBoost | 0.728 (0.721, 0.736) | 0.312 (0.300, 0.324) | −0.038 (−0.044, −0.032) | 0.002 |
| LACE logistic regression | 0.659 (0.652, 0.667) | 0.240 (0.230, 0.251) | −0.107 (−0.115, −0.100) | 0.002 |

**Abbreviations:** AUROC = area under the receiver operating characteristic curve; CI = confidence interval; LACE = Length of stay, Acuity of admission, Charlson Comorbidity Index, and Emergency department visits; PR-AUC = area under the precision-recall curve.

### Single-patient explanation

We illustrate an AToP explanation in **Figure 2** for a test patient with an oncologic trajectory spanning five admissions over 911 days. The patient was readmitted two days after the index discharge, with a predicted readmission probability of 0.93. The largest attributed contribution was a cross-visit pattern consisting of cancer pain at the second admission followed by co-occurring cancer anemia and cancer pain at the index admission (+0.83 of the total ΣIG of +0.97). The case was selected to illustrate the explanation rather than represent the cohort. Additional examples involving psychiatric, obstetric, and oncologic patients are presented in **Figures S2A–C**.

**Figure 2.**
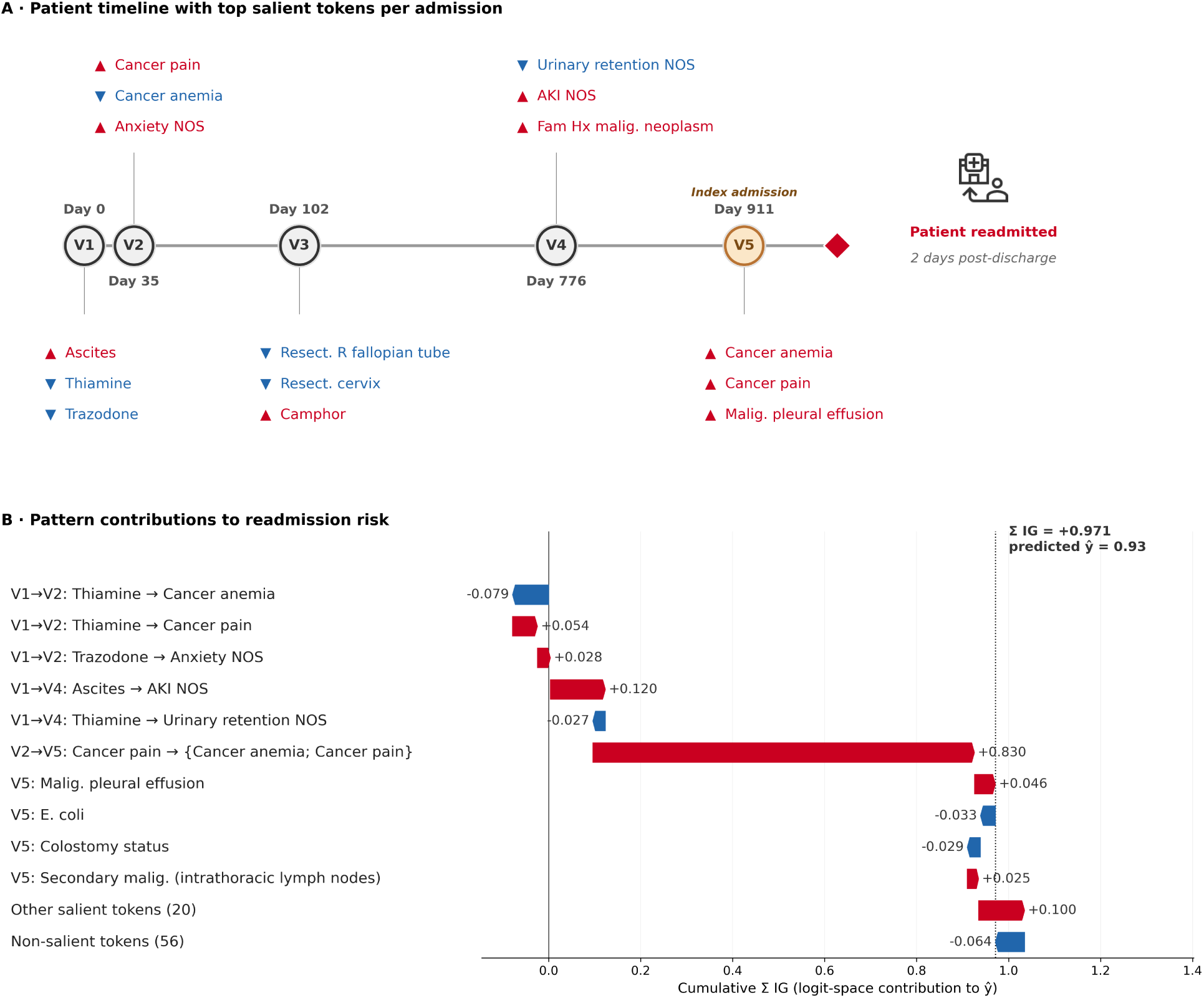
Single-patient AToP explanation. (A) Five-admission timeline with selected salient clinical events and the observed readmission. Red upward triangles indicate positive attribution; blue downward triangles indicate negative attribution. (B) Signed clinical-token attribution (ΣIG, logit scale) allocated to within-visit and cross-visit patterns, with remaining tokens shown separately. V labels identify the visits matched by each pattern. Each token position is assigned to at most one displayed pattern; the bars are attribution allocations, not independent pattern effects (Supplementary Methods). Clinical-token ΣIG = +0.971; attribution to [CLS] and [VISIT] = +1.310; reference logit = +0.302; residual = +0.058; patient logit = +2.641. NOS = not otherwise specified; AKI = acute kidney injury; Fam Hx = family history; malig. = malignant; Resect. = resection; R = right.

Because patterns can overlap, each salient token position is assigned to only one pattern in the attribution waterfall (**Figure 2B**), using a fixed priority rule (**Supplementary Methods**). The resulting contributions are allocations rather than independent pattern effects.

### Token-level global attribution

In **Figure 3**, we compare population-weighted and carrier-conditional token attribution in the test cohort (Equations 1–2). The population-weighted ranking (**Figure 3A**) was dominated by common codes with modest attribution among carriers, including MDD single episode NOS (not otherwise specified), live birth, and chest pain NOS. In contrast, the carrier-conditional ranking (**Figure 3B**) highlighted less prevalent codes with high attribution among carriers, including psychosis NOS, acetaminophen overdose/self-harm, severe MDD, and bipolar manic episode. Only 7 tokens appeared in both top-30 rankings.

**Figure 3.**
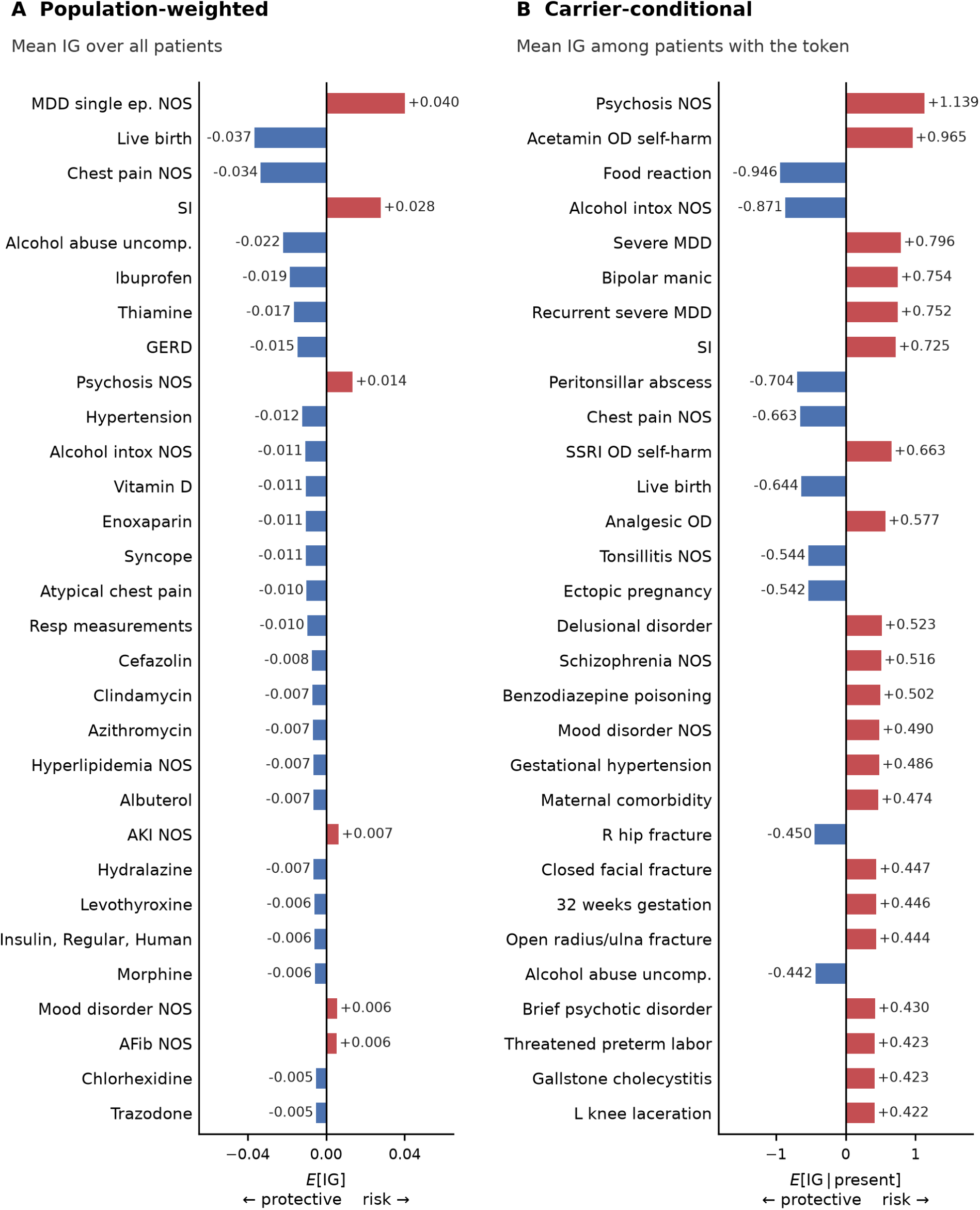
Token attribution at the population and carrier levels. The 30 tokens with the largest absolute mean signed IG attribution, calculated (A) across all test patients, with noncarriers contributing zero, and (B) among patients carrying each token. Red indicates positive attribution and blue negative attribution. Horizontal-axis scales differ between panels. MDD = major depressive disorder; ep. = episode; NOS = not otherwise specified; SI = suicidal ideation; uncomp. = uncomplicated; intox = intoxication; Resp = respiratory; GERD = gastroesophageal reflux disease; AKI = acute kidney injury; AFib = atrial fibrillation; OD = overdose; Acetamin = acetaminophen; SSRI = selective serotonin reuptake inhibitor; R = right; L = left.

### Pattern-level global attribution

Mining identified 70,278 distinct patterns in the training cohort (**Table S3**). After applying minimum carrier counts in the test cohort and Jaccard deduplication, 17,891 all-pattern and 9,213 cross-visit representatives remained for ranking. In **Figure 4**, we compare carrier-averaged ΣIG rankings across all pattern types (**Figure 4A**) and cross-visit patterns only (**Figure 4B**). The all-pattern ranking was dominated by within-visit co-occurrences, such as {Severe MDD; SI}. In contrast, the cross-visit ranking highlighted psychiatric, oncologic, and obstetric trajectories, including {MDD; SI} → {Recurrent severe MDD; SI} (**Tables 3–4**).

**Figure 4.**
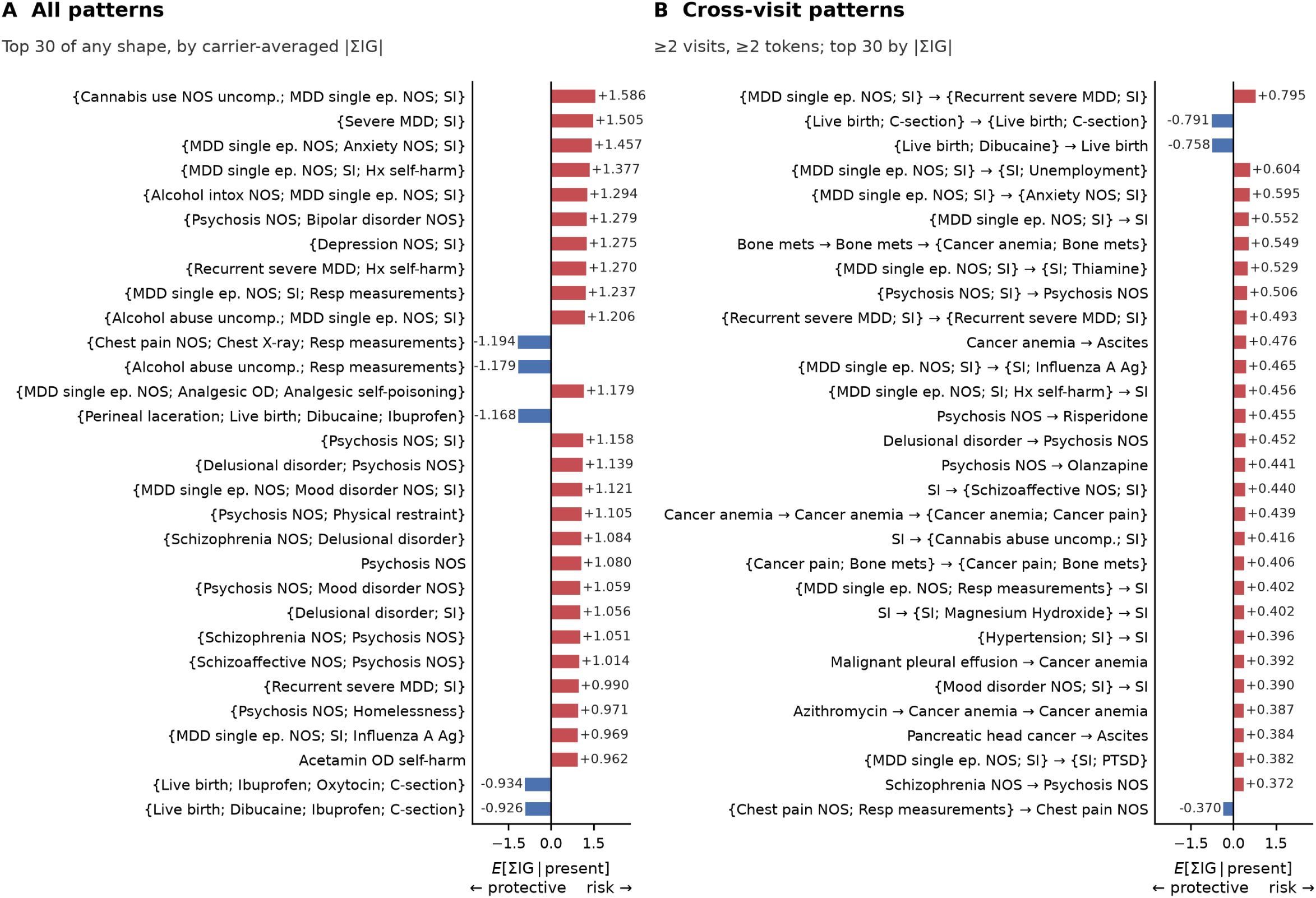
Attribution rankings for all patterns and cross-visit patterns. The 30 patterns with the largest absolute carrier-averaged ΣIG (mean sum of matched-token IG values among carriers) after Jaccard deduplication (threshold 0.20): (A) all pattern types; (B) sequences spanning at least two visits and two distinct tokens. Red indicates positive attribution and blue negative attribution. Both panels share a horizontal-axis scale. MDD = major depressive disorder; ep. = episode; NOS = not otherwise specified; SI = suicidal ideation; Hx = history of; uncomp. = uncomplicated; intox = intoxication; Resp = respiratory; OD = overdose; Acetamin = acetaminophen; C-section = cesarean delivery; Ag = antigen; PTSD = post-traumatic stress disorder; mets = metastases.

**Table 3.**
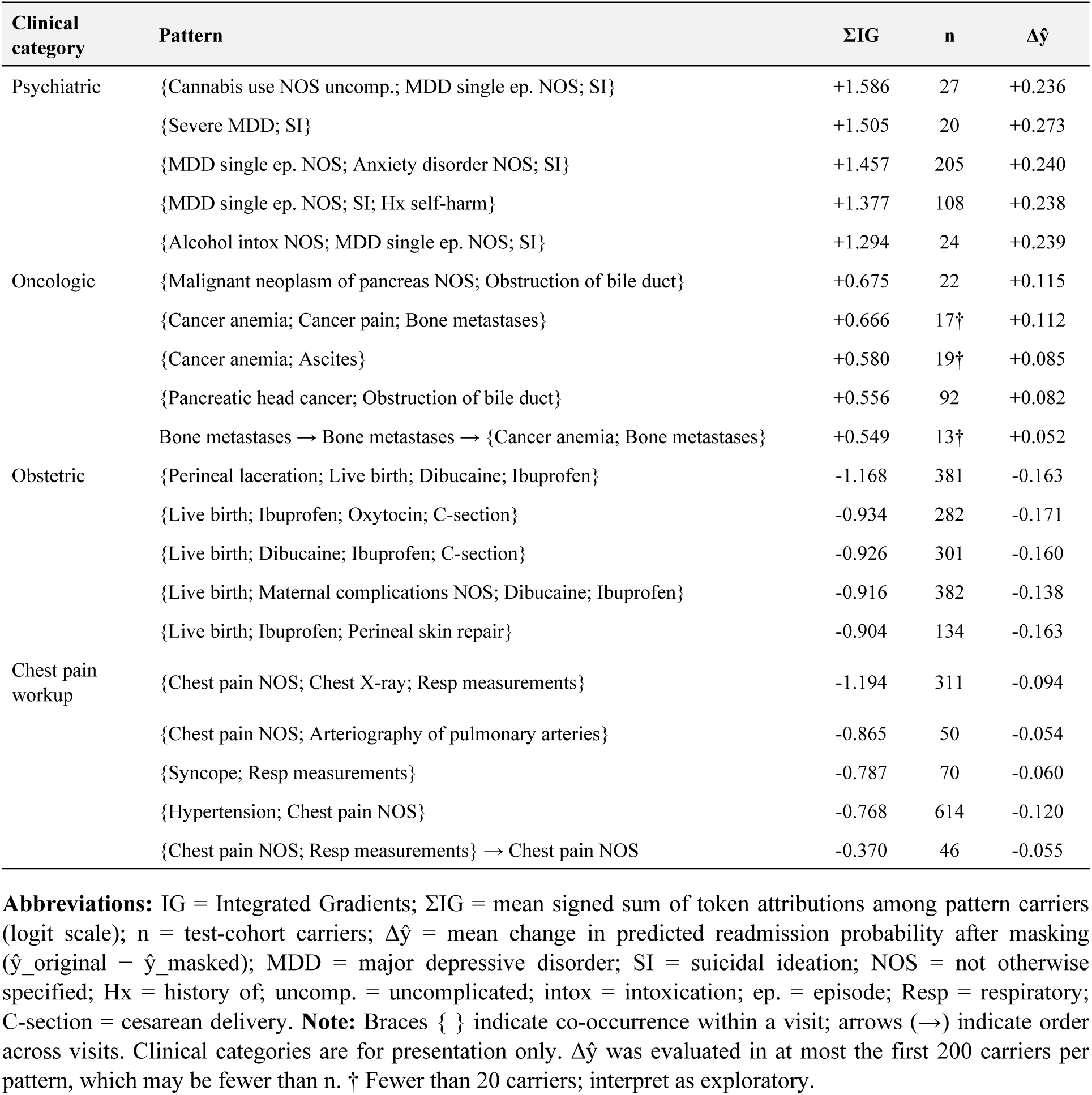
Top attribution patterns by clinical category.

| **Clinical category** | **Pattern** | **ΣIG** | **n** | **Δŷ** |
|---|---|---|---|---|
| Psychiatric | {Cannabis use NOS uncomp.; MDD single ep. NOS; SI} | +1.586 | 27 | +0.236 |
|  | {Severe MDD; SI} | +1.505 | 20 | +0.273 |
|  | {MDD single ep. NOS; Anxiety disorder NOS; SI} | +1.457 | 205 | +0.240 |
|  | {MDD single ep. NOS; SI; Hx self-harm} | +1.377 | 108 | +0.238 |
|  | {Alcohol intox NOS; MDD single ep. NOS; SI} | +1.294 | 24 | +0.239 |
| Oncologic | {Malignant neoplasm of pancreas NOS; Obstruction of bile duct} | +0.675 | 22 | +0.115 |
|  | {Cancer anemia; Cancer pain; Bone metastases} | +0.666 | 17† | +0.112 |
|  | {Cancer anemia; Ascites} | +0.580 | 19† | +0.085 |
|  | {Pancreatic head cancer; Obstruction of bile duct} | +0.556 | 92 | +0.082 |
|  | Bone metastases → Bone metastases → {Cancer anemia; Bone metastases} | +0.549 | 13† | +0.052 |
| Obstetric | {Perineal laceration; Live birth; Dibucaine; Ibuprofen} | -1.168 | 381 | -0.163 |
|  | {Live birth; Ibuprofen; Oxytocin; C-section} | -0.934 | 282 | -0.171 |
|  | {Live birth; Dibucaine; Ibuprofen; C-section} | -0.926 | 301 | -0.160 |
|  | {Live birth; Maternal complications NOS; Dibucaine; Ibuprofen} | -0.916 | 382 | -0.138 |
|  | {Live birth; Ibuprofen; Perineal skin repair} | -0.904 | 134 | -0.163 |
| Chest pain workup | {Chest pain NOS; Chest X-ray; Resp measurements} | -1.194 | 311 | -0.094 |
|  | {Chest pain NOS; Arteriography of pulmonary arteries} | -0.865 | 50 | -0.054 |
|  | {Syncope; Resp measurements} | -0.787 | 70 | -0.060 |
|  | {Hypertension; Chest pain NOS} | -0.768 | 614 | -0.120 |
|  | {Chest pain NOS; Resp measurements} → Chest pain NOS | -0.370 | 46 | -0.055 |

**Abbreviations:** IG = Integrated Gradients; ΣIG = mean signed sum of token attributions among pattern carriers (logit scale); n = test-cohort carriers; Δŷ = mean change in predicted readmission probability after masking (ŷ\_original − ŷ\_masked); MDD = major depressive disorder; SI = suicidal ideation; NOS = not otherwise specified; Hx = history of; uncomp. = uncomplicated; intox = intoxication; ep. = episode; Resp = respiratory; C-section = cesarean delivery. **Note:** Braces { } indicate co-occurrence within a visit; arrows (→) indicate order across visits. Clinical categories are for presentation only. Δŷ was evaluated in at most the first 200 carriers per pattern, which may be fewer than n. † Fewer than 20 carriers; interpret as exploratory.

**Table 4.** Top cross-visit attribution patterns by clinical category.

| **Clinical category** | **Pattern** | **ΣIG** | **n** | **Δŷ** |
|---|---|---|---|---|
| Psychiatric | {MDD single ep. NOS; SI} → {Recurrent severe MDD; SI} | +0.795 | 36 | +0.061 |
|  | {MDD single ep. NOS; SI} → {SI; Unemployment NOS} | +0.604 | 10† | +0.054 |
|  | {MDD single ep. NOS; SI} → {Anxiety disorder NOS; SI} | +0.595 | 37 | +0.041 |
|  | {MDD single ep. NOS; SI} → SI | +0.552 | 201 | +0.041 |
|  | {MDD single ep. NOS; SI} → {SI; Thiamine} | +0.529 | 29 | +0.032 |
| Oncologic | Bone metastases → Bone metastases → {Cancer anemia; Bone metastases} | +0.549 | 13† | +0.052 |
|  | Cancer anemia → Ascites | +0.476 | 21 | +0.060 |
|  | Cancer anemia → Cancer anemia → {Cancer anemia; Cancer pain} | +0.439 | 21 | +0.044 |
|  | {Cancer pain; Bone metastases} → {Cancer pain; Bone metastases} | +0.406 | 16† | +0.055 |
|  | Malignant pleural effusion → Cancer anemia | +0.392 | 14† | +0.048 |
| Obstetric | {Live birth; C-section} → {Live birth; C-section} | -0.791 | 51 | -0.140 |
|  | {Live birth; Dibucaine} → Live birth | -0.758 | 165 | -0.137 |
|  | {Live birth; Maternal complications NOS} → Maternal complications NOS | -0.351 | 23 | -0.061 |
|  | Live birth → Influenza A Ag | -0.284 | 105 | -0.046 |
|  | Oxazepam → Maternal comorbidity | +0.252 | 25 | +0.034 |
| Chest pain workup | {Chest pain NOS; Resp measurements} → Chest pain NOS | -0.370 | 46 | -0.055 |
|  | Chest pain NOS → Simvastatin | -0.269 | 73 | -0.049 |
|  | Chest pain NOS → Chest pain NOS → Atypical chest pain | -0.256 | 35 | -0.045 |
|  | Chest pain NOS → Clindamycin | -0.251 | 39 | -0.051 |
|  | Chest pain NOS → Chest pain NOS → GERD | -0.232 | 32 | -0.040 |
| Hepatobiliary | {Obstruction of bile duct; ERCP biliary stent} → Obstruction of bile duct | +0.270 | 57 | +0.036 |
|  | Obstruction of bile duct → Percutaneous biliary drainage | +0.254 | 17† | +0.029 |
|  | {GERD; Obstruction of bile duct} → Obstruction of bile duct | +0.158 | 28 | +0.017 |
|  | Acute pancreatitis NOS → Other chronic pain → Other chronic pain | -0.152 | 24 | -0.027 |
|  | Obstruction of bile duct → Intrahepatic bile duct carcinoma | +0.146 | 22 | +0.010 |
| Other | CVC placement → Venous catheterization, NEC → CVC placement → CVC placement | +0.236 | 19† | +0.024 |
|  | CVC placement → {Thiamine; CVC placement} → CVC placement | +0.228 | 7† | +0.016 |
|  | {Thiamine; Vitamin D} → Clindamycin | -0.217 | 27 | -0.040 |
|  | {Meropenem; CVC placement} → CVC placement → CVC placement | +0.217 | 13† | +0.027 |
|  | Lactulose → CVC placement → CVC placement → CVC placement | +0.206 | 24 | +0.022 |

**Abbreviations:** Additional abbreviations: CVC = central venous catheter; ERCP = endoscopic retrograde cholangiopancreatography; GERD = gastroesophageal reflux disease; NEC = not elsewhere classified; Ag = antigen. Other definitions and conventions are as in Table 3.

The all-pattern and cross-visit rankings shared only one pattern among their top 50. Jaccard deduplication reduced near-duplicate patterns and increased the diversity of clinical categories represented in the cross-visit ranking. Within-visit co-occurrences continued to dominate the all-pattern ranking across deduplication thresholds (**Figures S3–S4**). Within the all-pattern pool, population-weighted and carrier-averaged ΣIG rankings shared only 6 of their top 30 patterns (**Figure S5**).

### Clinical pattern categories and masking effects

In **Tables 3 and 4**, we present the top five patterns by absolute carrier-averaged ΣIG within each clinical category, for all patterns and cross-visit patterns, respectively. Masking the tokens in each displayed pattern changed predicted readmission probability in the direction indicated by its attribution (**Figure 5**). The same directional agreement was observed across all patterns tested in the broader masking analysis (**Figure S6**), with carrier-averaged ΣIG strongly correlated with mean masking effects (Spearman ρ = 0.94 overall, 0.79 for risk-increasing patterns, and 0.92 for protective patterns).

**Figure 5.**
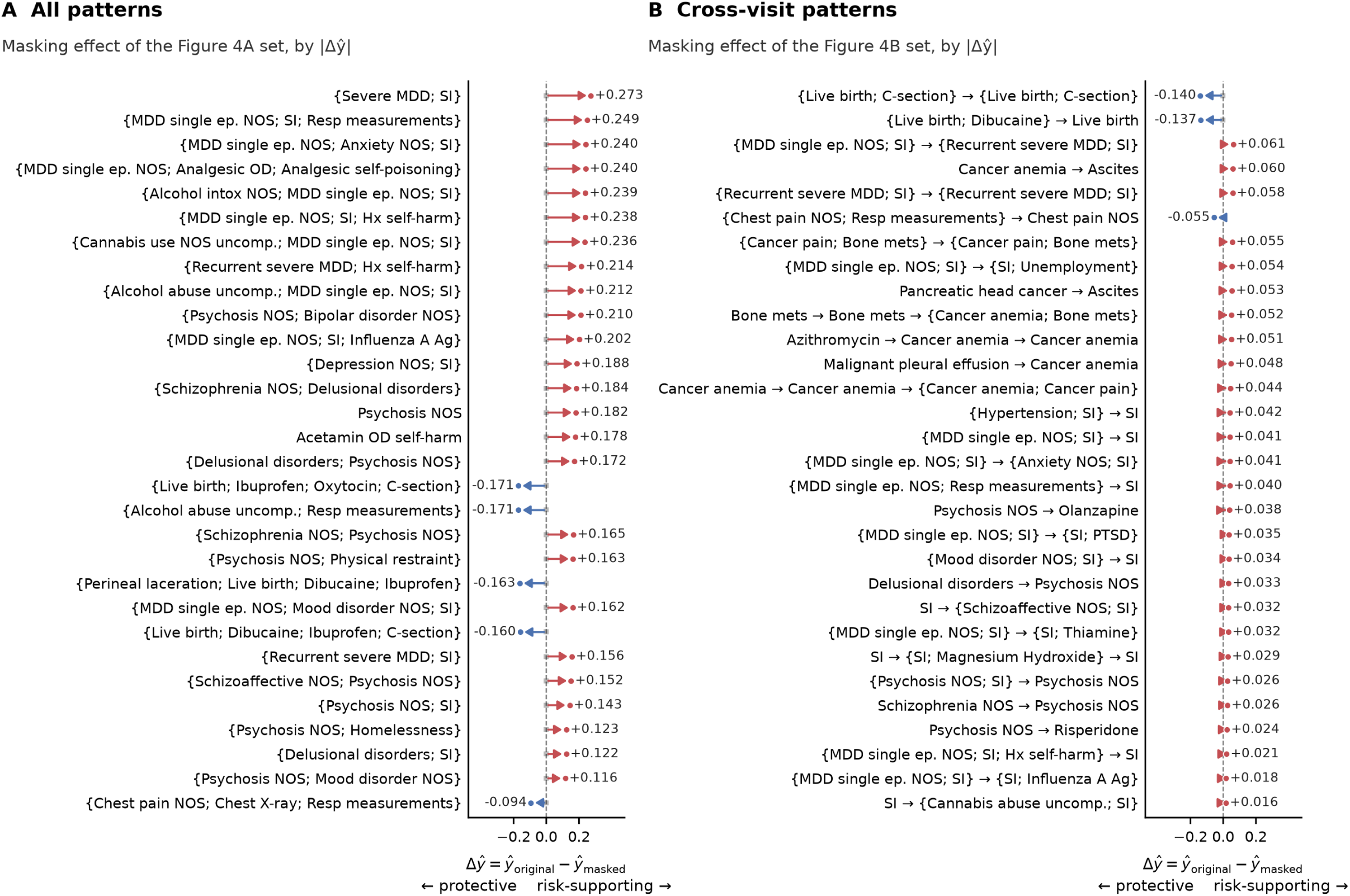
Masking effects for top-ranked patterns. Prediction changes after masking constituent tokens of the 30 highest-ranked (A) all-pattern and (B) cross-visit representatives in Figure 4. Δŷ = predicted probability before minus after masking, averaged over at most the first 200 test-cohort carriers per pattern. Positive values (red) indicate that masking lowered predicted risk; negative values (blue) indicate that masking raised it. Patterns are sorted by |Δŷ| within each panel. Both panels share a horizontal-axis scale. Abbreviations as in Figure 4.

To examine whether pattern attributions aligned with observed readmission risk, we estimated odds ratios for the 30 reported cross-visit patterns, adjusting for the number of admissions. The direction of model attribution agreed with the direction of the odds ratio for 25 of these patterns (**Table S4**).

### Token-level attribution faithfulness

To assess whether highly attributed tokens had greater influence on model predictions, we compared IG-ranked tokens with randomly selected salient tokens. Masking the five highest-ranked tokens produced a larger mean absolute change in predicted readmission probability than masking five randomly selected salient tokens (0.079 vs 0.056; Wilcoxon signed-rank p < 0.001).

Across selections of 1 to 10 tokens, masking the highest-attributed tokens produced larger prediction changes than masking randomly selected salient tokens, and retaining them better preserved the original predictions, with non-overlapping 95% bootstrap intervals at every k (**Figure S7**).

### Temporal-order analysis

For each cross-visit pattern, we compared attribution between two groups of patients: those whose records contained the pattern in its reported order and those whose records contained the same events in reverse order. Three of the 30 patterns in **Table 4** read the same in both directions and were excluded. For 26 of the remaining 27 patterns (96%), attribution magnitude was lower in the reversed-order group, with a mean attenuation of 30% (**Table S5**). The effect was weaker in a broader set of 1,714 patterns. Attribution magnitude was lower in the reversed-order group for 54% of them, and attribution direction was the same in both groups for 95% (**Figure S8**). Because the two groups contained different patients, these differences may also reflect patient characteristics rather than event order alone.

In the within-patient analysis, reversing the order of matched visits reduced attribution magnitude for 21 of the 27 patterns, with a mean attenuation of 14%. Among 722 patients, 59% showed a reduction in mean attribution magnitude, with a significant overall difference (one-sided Wilcoxon signed-rank p < 0.001; **Table S6**).

### Cross-method comparison

We compared Integrated Gradients (IG) with KernelSHAP at the patient, token, and pattern levels as an exploratory cross-implementation comparison. Per-patient attribution agreement is shown in **Figure S9**. The methods shared 25 of their top 30 carrier-conditional tokens (**Figure S10**). At the pattern level, they shared 21 of the top 30 all-pattern representatives but only 7 of the top 30 cross-visit representatives (**Figure S11**).

## Discussion

This study introduces AToP, an explainability framework that combines token attribution with temporal pattern mining to explain Transformer predictions from longitudinal EHR data. AToP extends conventional feature rankings by identifying combinations of clinical events within visits and sequences across visits, and by summarizing their attribution at the patient and cohort levels. In our readmission application, carrier-conditional attribution, defined as the mean attribution among patients whose records contain a given token, highlighted less prevalent clinical features that were obscured in population-weighted rankings. Separating cross-visit patterns from within-visit combinations revealed psychiatric and oncologic trajectories that were less prominent in the all-pattern rankings.

Masking tokens in patterns with positive attribution lowered predicted readmission probabilities, whereas masking those with negative attribution raised them. These findings support correspondence between attribution and model behavior. Temporal-order analyses provided more qualified evidence: attribution magnitudes decreased under reversal for most reported patterns, but only about half of the broader pattern pool. IG and KernelSHAP showed substantial agreement on token rankings but limited agreement on cross-visit pattern rankings, consistent with documented disagreement between explanation methods.^33^

Previous approaches have represented temporal information in different ways. Attention-based models such as RETAIN assign importance weights to visits and clinical features, although the faithfulness of attention as an explanation remains debated.^13–15^ Other work has mined sequential patterns before applying SHAP to explain Transformer predictions.^24^ AToP reverses this process by using attribution to guide pattern discovery and then scoring the identified patterns among patients whose records contain them.

Many highly attributed patterns included conditions already known to be associated with readmission. Patterns that increased predicted risk were mainly psychiatric sequences involving major depressive disorder and suicidal ideation, and oncologic sequences involving cancer anemia, bone metastases, and ascites. Both have established links to readmission: depressive symptoms are associated with higher readmission rates after medical hospitalization,^34^ and patients with advanced cancer are frequently rehospitalized.^35^ Patterns that decreased predicted risk were mainly obstetric patterns and chest pain evaluations. For obstetric patterns, this is consistent with national data showing lower readmission rates after childbirth than after admissions for mental health conditions or cancer.^36^ AToP thus recovered recognizable clinical risk profiles rather than new patient groups. Its added value lies in showing how combinations and sequences of these conditions contribute to the model’s predictions.

Unspecified diagnosis codes appeared frequently among highly attributed tokens and patterns, including psychosis NOS, mood disorder NOS, and recurrent sequences involving depression and suicidal ideation. This may reflect diagnostic uncertainty, documentation practices, or coding conventions. Other patterns combined symptoms with diagnostic procedures, such as chest pain with chest radiography and respiratory measurements. AToP therefore surfaces care processes as well as clinical conditions, and distinguishing the two requires pattern-specific studies to replicate the findings and assess their plausibility.

### Limitations

Our evaluation assessed whether AToP explanations reflected model behavior, not whether they improved clinical decisions.^37^ Pattern attribution sums constituent token attributions and does not isolate the independent effects of their combination or temporal order. The explanations also exclude attribution to [CLS] and [VISIT] tokens and do not separately quantify the contribution of admission count.

Masking may create inputs that differ from those encountered during training.^38^ Because the masked patterns were selected using IG, this analysis does not establish whether they are the most influential patterns among all candidates. Between-patient reversal may reflect differences in patient characteristics, while within-patient reversal changes entire visits, including their other events and recency.

IG and KernelSHAP differed in reference distribution, output scale, sparsity, and pattern scoring. KernelSHAP also reused representatives selected through IG-based Jaccard deduplication, so the limited cross-visit agreement cannot be attributed solely to the attribution methods. The 80% saliency threshold was heuristic. Risk-increasing pattern rankings were relatively stable across evaluated thresholds with the mined dictionary fixed, but protective rankings were less stable (**Figure S12**). Patterns with fewer than 20 carriers should be considered exploratory.

Evaluation was limited to one dataset, task, and model architecture. Applying AToP to larger pretrained models, including EHR foundation models, is a natural next step. Single-admission patients were labeled as not readmitted by construction, readmissions outside MIMIC-IV may have been missed, and death was not modeled as a competing outcome. The performance difference from the baselines reflects both feature availability and model architecture.

### Conclusion

This work introduced AToP, an explainability framework that organizes token attributions from a Transformer into temporal patterns in longitudinal EHR data. By identifying combinations of clinical events within visits and sequences across visits, AToP provides explanations of model predictions that go beyond conventional rankings of individual features.

## Author Contributions

AB conceived the study, developed the methods and software, performed the analyses, and drafted the manuscript. HK conceived the study, supervised the work, and revised the manuscript. Both authors approved the final version and are accountable for the work.

## Funding

Research reported in this publication was supported by the National Library of Medicine (NLM) of the National Institutes of Health under Award Number T15LM013979. The content is solely the authors’ responsibility and does not necessarily represent the official views of the National Institutes of Health.

## Conflict of Interest

The authors declare no competing interests relevant to this work.

## Ethics Statement

This study used the de-identified MIMIC-IV v3.1 database, accessed under the PhysioNet credentialed data use agreement after completion of the required CITI training. Analysis of this publicly available de-identified resource did not require separate institutional review board approval.

## Data Availability

MIMIC-IV v3.1 is available to credentialed users through PhysioNet (https://physionet.org/content/mimiciv/3.1/). The AToP framework code is publicly available at https://github.com/belouali/atop. All analytic outputs referenced in the manuscript can be regenerated from MIMIC-IV v3.1 using the scripts in the repository.

## Supplementary Material

## Supplementary Methods

### Integrated Gradients reference baseline

We computed Integrated Gradients (IG) on the model’s pre-sigmoid logit using an all-[PAD] embedding reference and 30 integration steps. The reference retained each patient’s padding mask and position embeddings, so its output varied across patients. Completeness was assessed by comparing the sum of attributions with the difference between the patient’s logit and the reference logit. The completeness residual had a median absolute value of 0.012 logits (90th percentile 0.032), or 1.8% of the logit difference. Across 300 sampled test patients, the [CLS] and [VISIT] positions carried a median 46% of total absolute attribution.

Patient-level explanations and pattern analyses used attribution to clinical tokens only, excluding [CLS] and [VISIT] positions. The full attribution accounting, including the reference logit and special-token contributions, is provided for the examples in **Figures 2 and S2A–C**.

### Pattern scoring and position allocation

For global rankings, patterns were matched to patients’ complete records in chronological order. Each pattern step was assigned to the earliest eligible visit after the preceding step, with all tokens in a step required to occur in the same visit. For each carrier, we summed the signed IG values of the matched tokens, using their first occurrence within each assigned visit. Scores were averaged across carriers to obtain carrier-averaged ΣIG. Patterns were scored independently, including when they shared token positions.

For patient-level explanations (**Figure 2**), patterns were matched to salient visit blocks. Matches were prioritized by earliest matched visit, then decreasing number of visit blocks, and finally decreasing |ΣIG|. Each salient token position was assigned to at most one pattern, with remaining attribution displayed separately.

### Clinical pattern categorization

Patterns were assigned to clinical categories using ordered keyword matching on their readable labels. Categories were evaluated in the following order: psychiatric, oncologic, hepatobiliary, obstetric, substance use, chest pain workup, and other. The first matching category was assigned. These categories were used only for presentation and did not influence pattern mining, attribution, or ranking. The keyword lists and assignment rules are available in the AToP repository (atop/explain/clinical_categories.py).

### Preprocessing and vocabulary composition

ICD-9 diagnosis and procedure codes were mapped to ICD-10 using the CMS General Equivalence Mappings.^28^ Only unambiguous one-to-one mappings were retained; other codes remained in ICD-9. Chronic conditions, identified using a fixed ICD-prefix list, were retained at their first occurrence or when recorded as the primary diagnosis. Acute diagnoses were retained at every occurrence.

Prescription drugs were mapped to RxNorm ingredient concepts using the RxNav API, first by National Drug Code (NDC) and then by drug name when an NDC match was unavailable. Name-based matching used the first word of the order name. Each ingredient was retained at its first recorded prescription. Ingredients prescribed in more than 25% of admissions, ten near-universal ingredients, and a fixed list of administrative and screening codes were excluded.

Tokens occurring fewer than twice in the training set were mapped to [PAD]. The final vocabulary contained 24,005 tokens: 14,960 diagnoses, 7,877 procedures, 1,165 RxNorm ingredients, and three special tokens ([VISIT], [CLS], and [PAD]). The code lists and mapping procedures are available in the repository.

### Outcome label

A readmission was a subsequent admission beginning within 0 to 30 days after index discharge, including the day of discharge. Admissions were not merged, and no minimum interval was imposed. Admissions ending in in-hospital death were excluded before labeling. Deaths after discharge were not included in outcome ascertainment.

### Test-set use in deduplication

Patterns were mined exclusively from the training cohort. Test-cohort carrier sets were used for Jaccard deduplication, and test-cohort attributions determined which representatives were retained for reporting. Neither procedure used test outcomes or altered model training or pattern discovery.

### Validation settings

#### Pattern masking and within-patient reversal

Masking included the patterns displayed in **Tables 3 and 4** and additional highly ranked patterns, yielding 266 unique patterns. For each pattern, up to 200 test-cohort carriers were evaluated in cohort order. Within-patient reversal used up to 80 carriers per displayed cross-visit pattern, also selected in cohort order.

#### Token-level faithfulness

We randomly sampled 500 test patients with at least three salient tokens. Comprehensiveness and sufficiency were evaluated for the top 1 to 10 tokens, including patients with at least the corresponding number of salient tokens at each step. The random baseline used five draws per patient. Confidence intervals were calculated using 1,000 patient-level bootstrap resamples (seed 42).

#### KernelSHAP comparison

KernelSHAP treated each patient’s non-special tokens as features and replaced masked tokens with [PAD]. The background consisted of ten randomly generated masks per patient, retaining each token with probability 0.5. Approximately 50 coalition samples were used per patient across the analytic test cohort (n = 43,602), and 200 were used for the per-patient comparison in **Figure S9**.

KernelSHAP explained predicted probabilities, whereas IG explained logits. The SHAP implementation used default L1 regularization, retaining at most ten nonzero feature attributions per patient. SHAP pattern scores summed attribution for unique constituent tokens, whereas IG used matched token positions. KernelSHAP rankings reused the IG-selected Jaccard representatives rather than independently selecting representatives. Extreme SHAP values (absolute value greater than 10) were excluded from token summaries and affected pattern scores. Analyses used shap version 0.49.1.

#### Temporal-order comparison

The between-patient analysis included reported cross-visit patterns and additional multi-visit patterns with at least 100 test-cohort carriers. Of 3,447 candidate patterns, 1,714 had at least 20 patients matching each ordering and were included in the broader comparison (**Figure S8**). Both orderings were matched using attribution-salient visit blocks rather than complete records, accounting for the different carrier counts in **Tables 4 and S5**.

#### Attribution direction and observed readmission

We examined associations between the reported cross-visit patterns and observed readmission among test patients with at least two admissions. For each pattern, a separate logistic regression modeled readmission using pattern presence and the log-transformed number of admissions as predictors. Adjusted odds ratios and 95% Wald confidence intervals are reported in **Table S4**.

---

**Figure S1.**
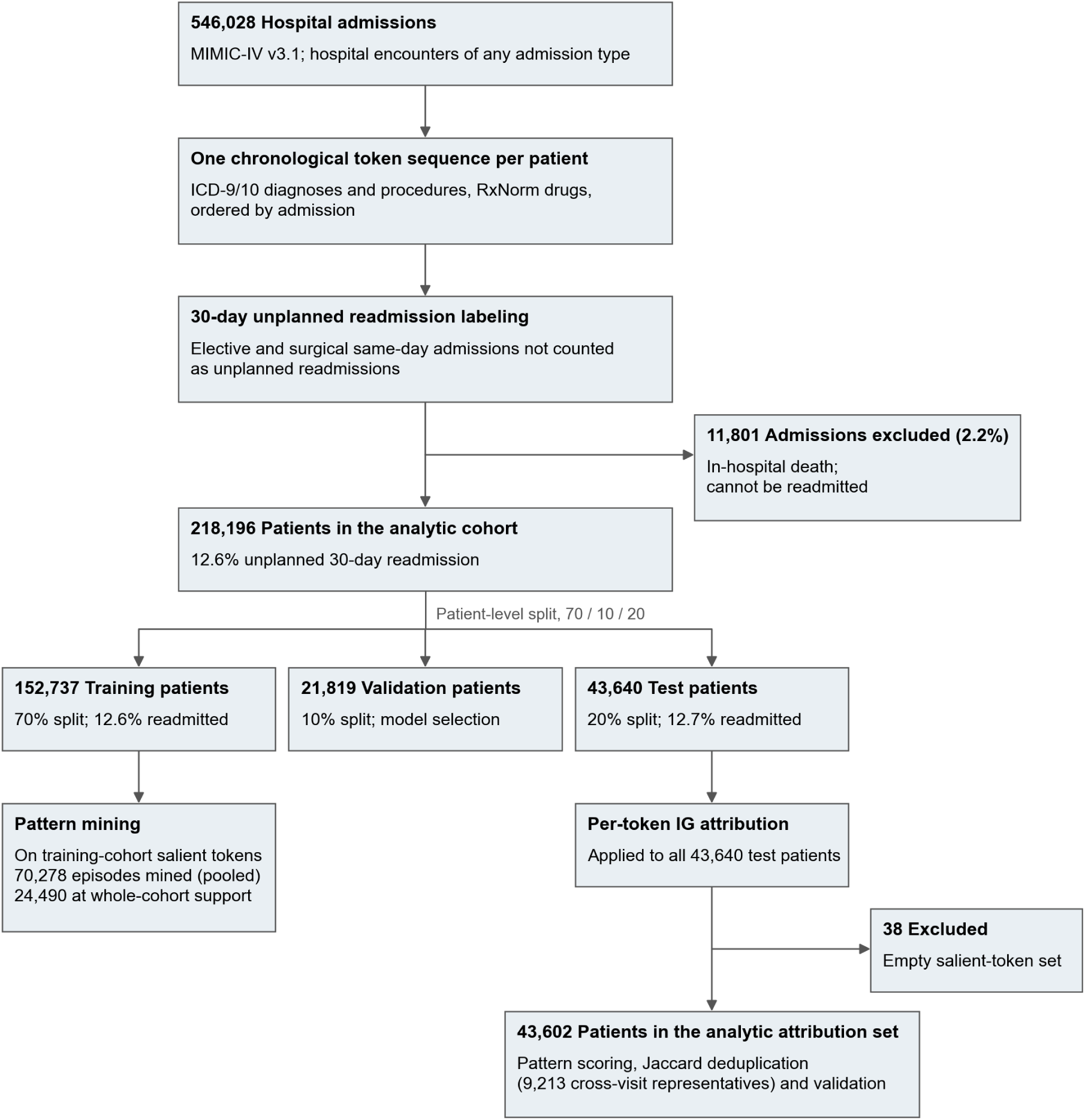
Cohort and analysis flowchart. Derivation of the 218,196-patient cohort from MIMIC-IV v3.1, including exclusions, unplanned 30-day readmission labels, and the training, validation, and test splits. The test cohort contained 43,640 patients; 43,602 had nonempty salient-token sets and were included in AToP attribution analyses. Training-cohort pattern mining and test-cohort attribution, carrier matching, and deduplication are shown separately.

**Figure S2A.**
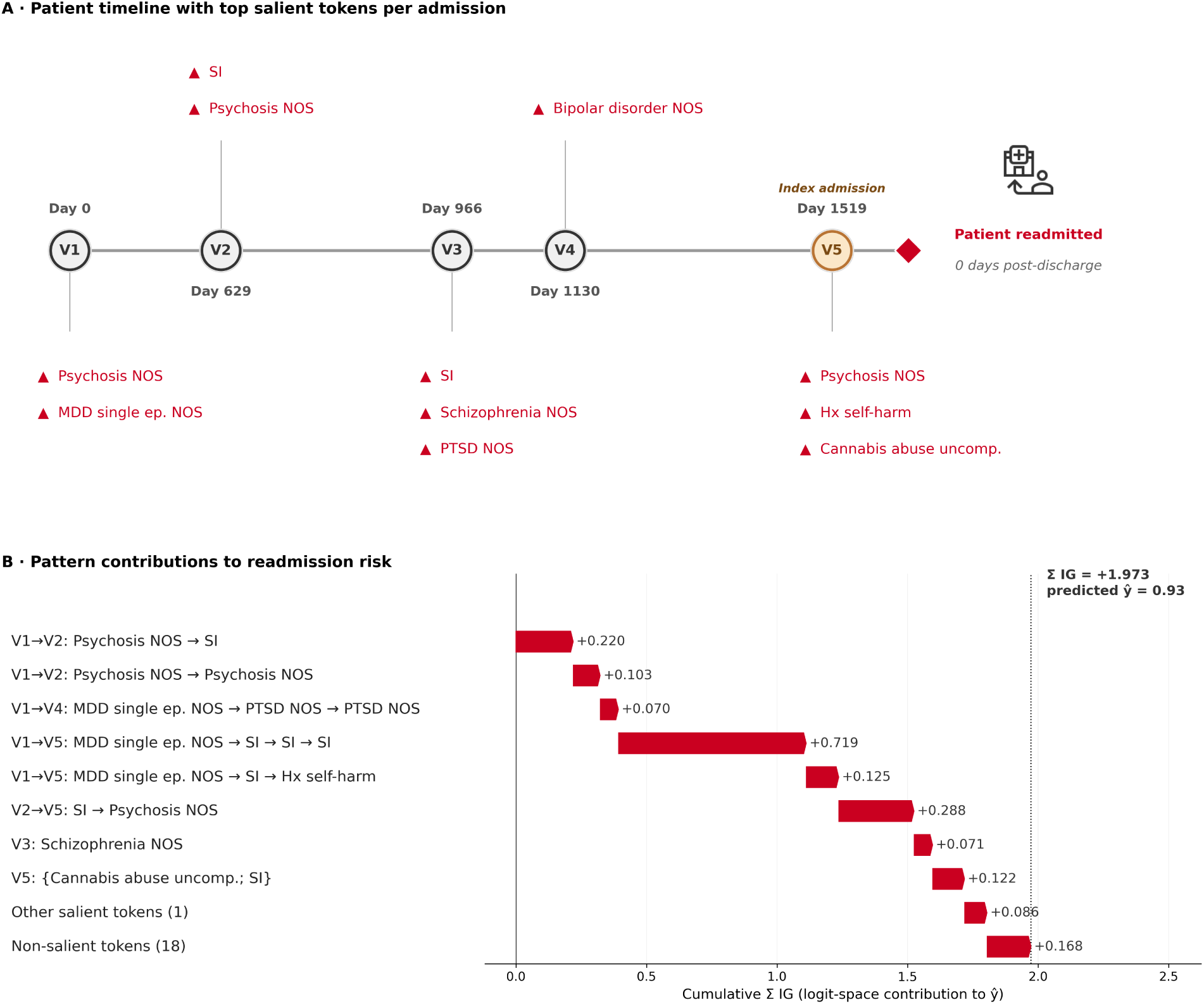
Additional patient explanation: psychiatric true positive. Patient with recurrent psychosis NOS across admissions, shown as an admission timeline (A) and allocated pattern attributions (B), as in Figure 2. Clinical-token ΣIG = +1.973; attribution to [CLS] and [VISIT] = +0.733; reference logit = −0.220; residual = +0.046; patient logit = +2.533. NOS = not otherwise specified.

**Figure S2B.**
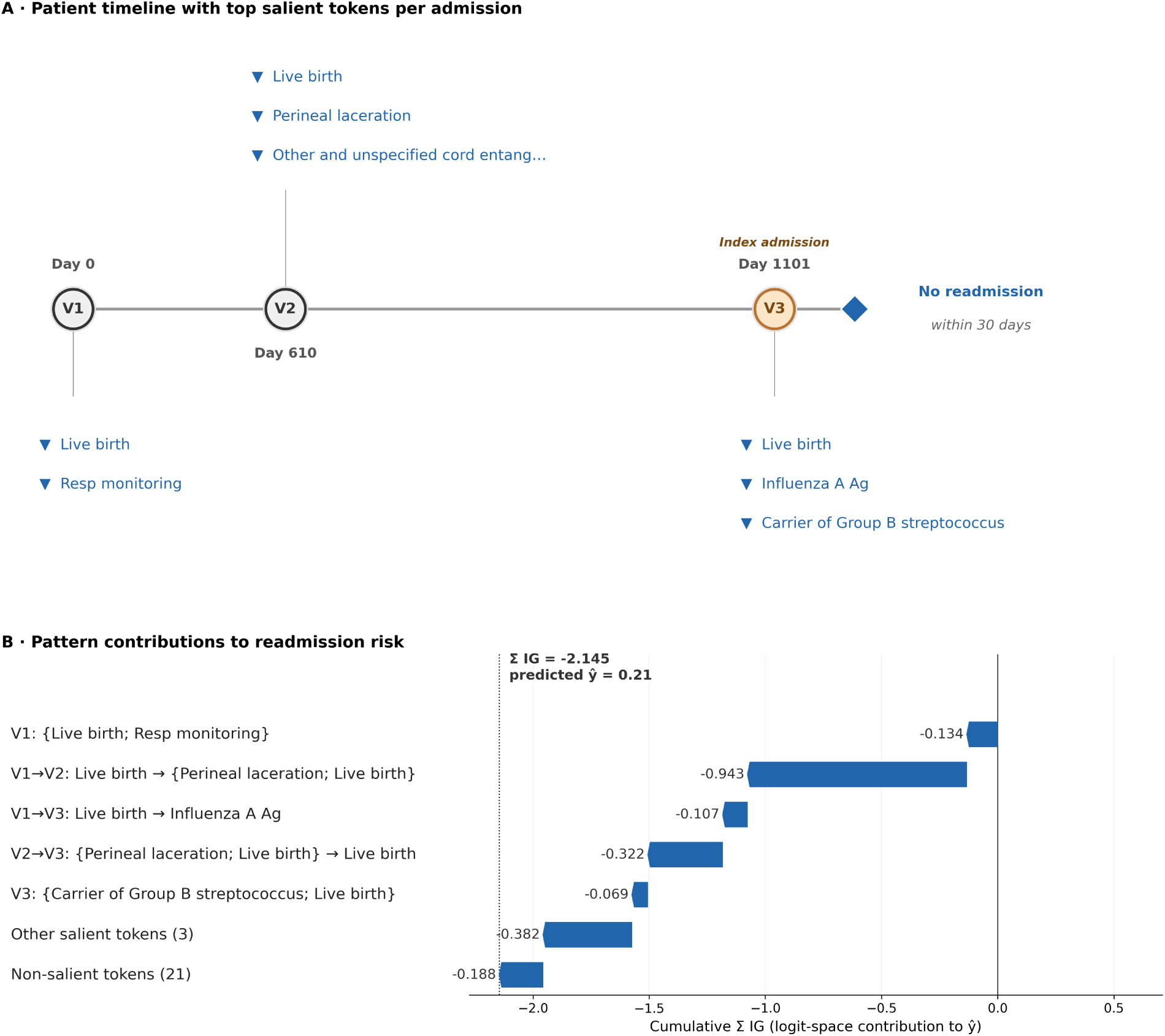
Additional patient explanation: obstetric true negative. Patient with an obstetric presentation involving delivery and laceration repair, shown as an admission timeline (A) and allocated pattern attributions (B), as in Figure 2. Clinical-token ΣIG = −2.145; attribution to [CLS] and [VISIT] = +1.062; reference logit = −0.198; residual = −0.019; patient logit = −1.300.

**Figure S2C.**
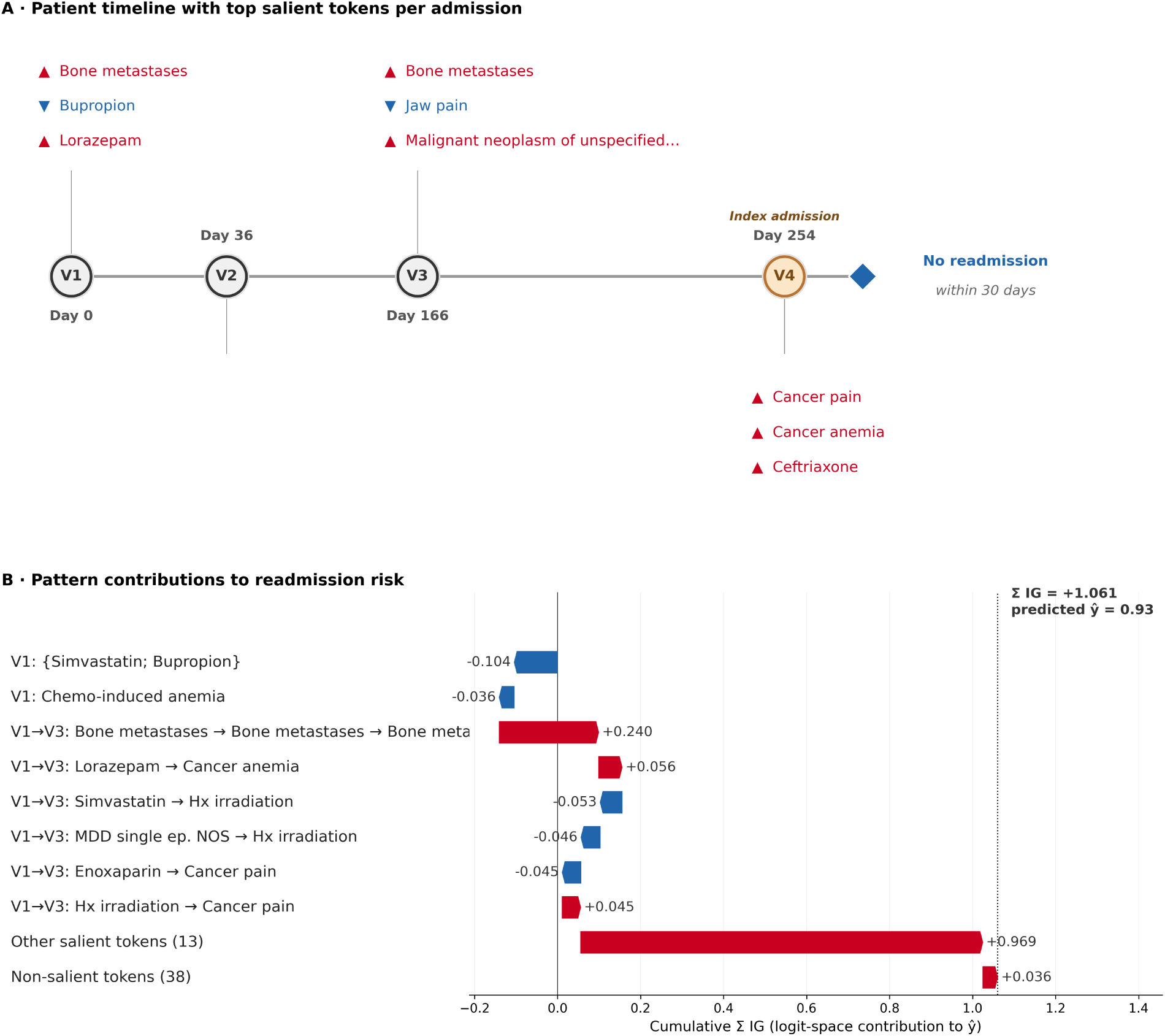
Additional patient explanation: oncology false positive. Patient with metastatic cancer whose predicted readmission probability was 0.93 but who was not readmitted within 30 days. The admission timeline (A) and attribution allocation (B) include a repeated bone-metastases pattern (+0.240 ΣIG) and other salient tokens (+0.969 ΣIG), as in Figure 2. Clinical-token ΣIG = +1.061; attribution to [CLS] and [VISIT] = +1.280; reference logit = +0.145; residual = +0.048; patient logit = +2.534.

**Figure S3.**
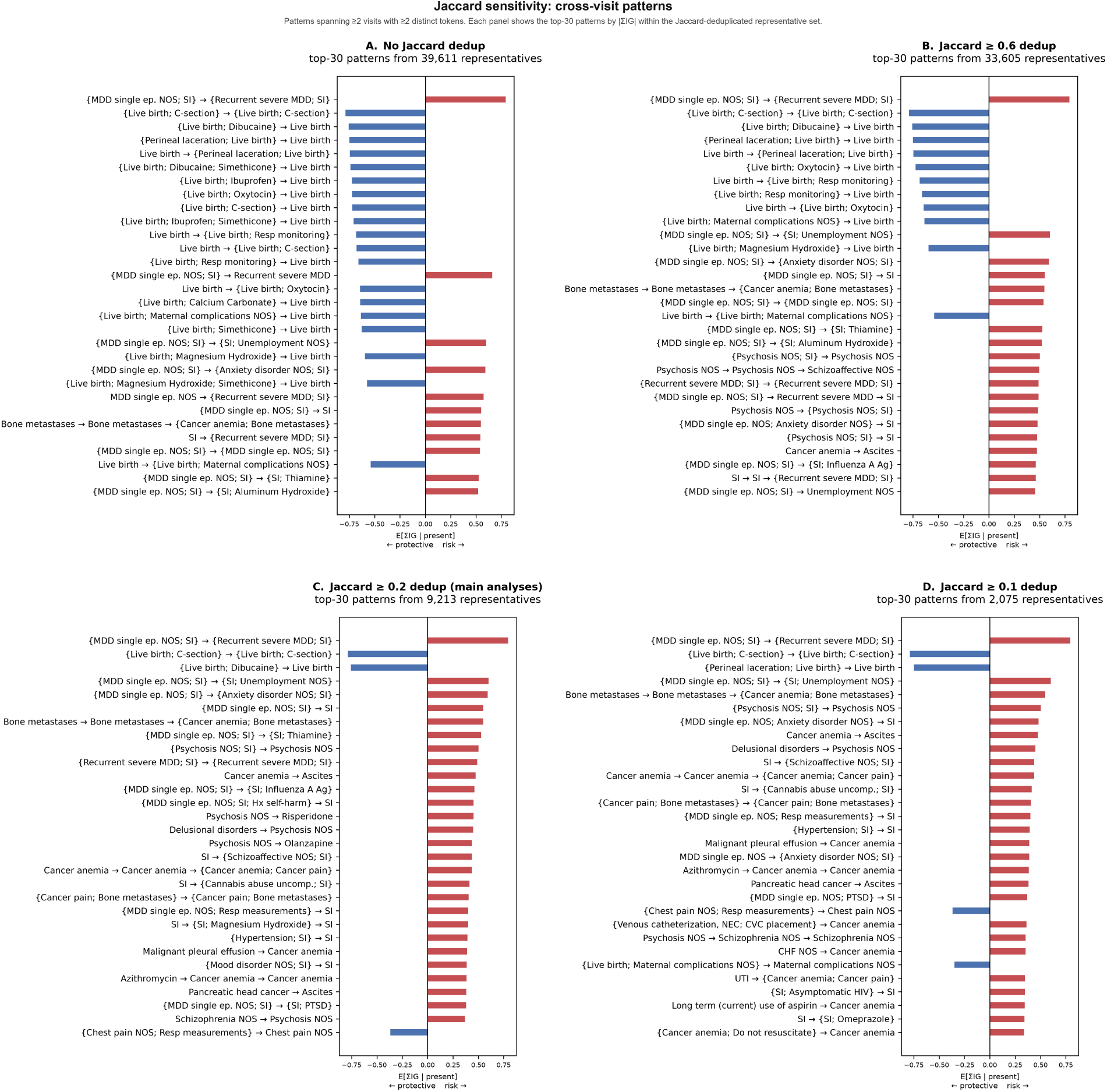
Sensitivity of cross-visit rankings to Jaccard deduplication. Top 30 cross-visit patterns by absolute carrier-averaged ΣIG with (A) no deduplication (39,611 representatives), (B) threshold 0.60 (33,605), (C) threshold 0.20 (9,213; main analysis), and (D) threshold 0.10 (2,075). Lower thresholds group more patterns with overlapping test-cohort carrier sets; panels A and C share 8 of their top 30 patterns.

**Figure S4.**
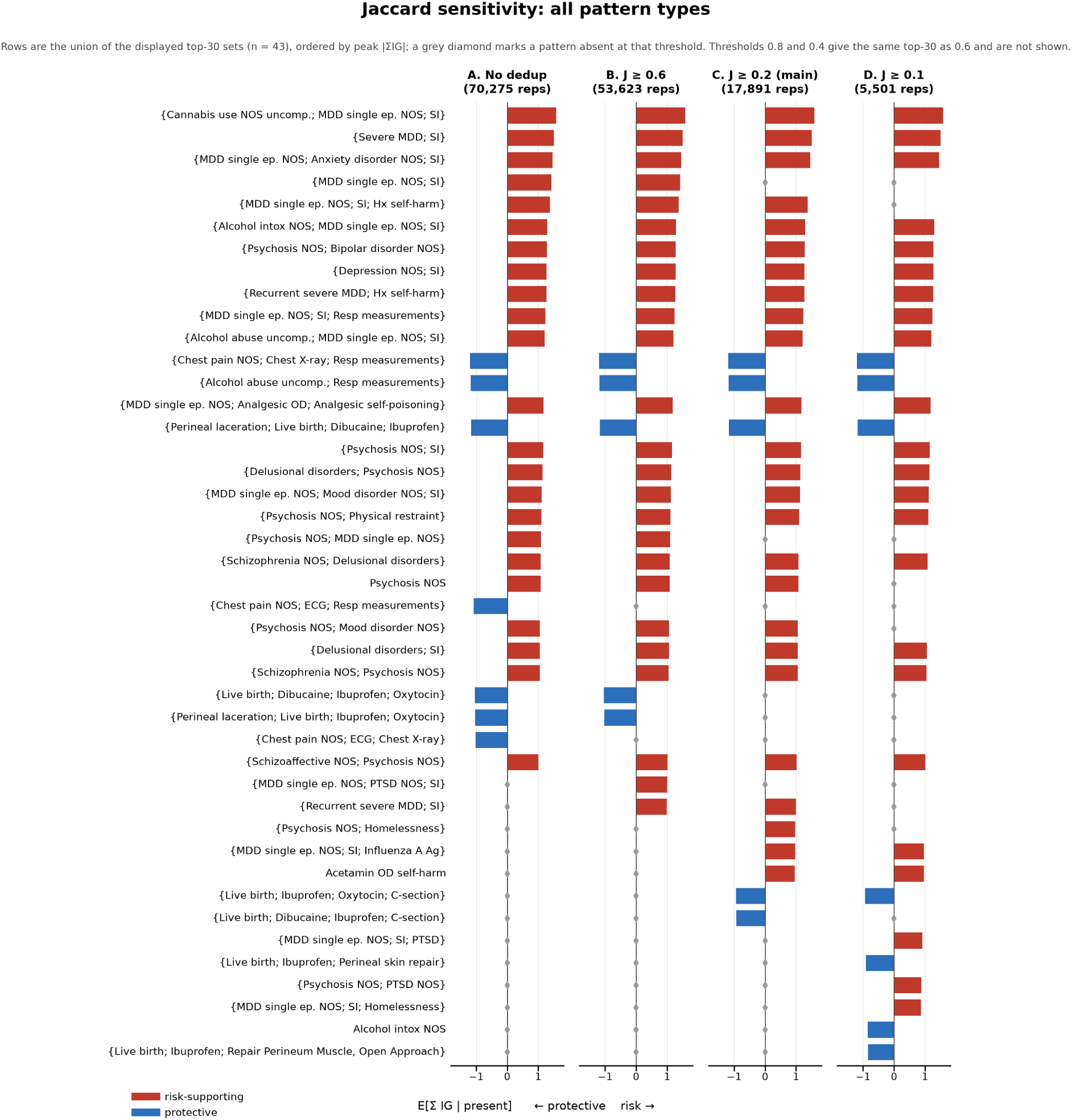
Sensitivity of all-pattern rankings to Jaccard deduplication. Top 30 patterns across all types at (A) no deduplication (70,275 representatives), (B) threshold 0.60 (53,623), (C) threshold 0.20 (17,891; main analysis), and (D) threshold 0.10 (5,501). Patterns are aligned across panels; a gray diamond indicates absence from that panel’s top 30.

**Figure S5.**
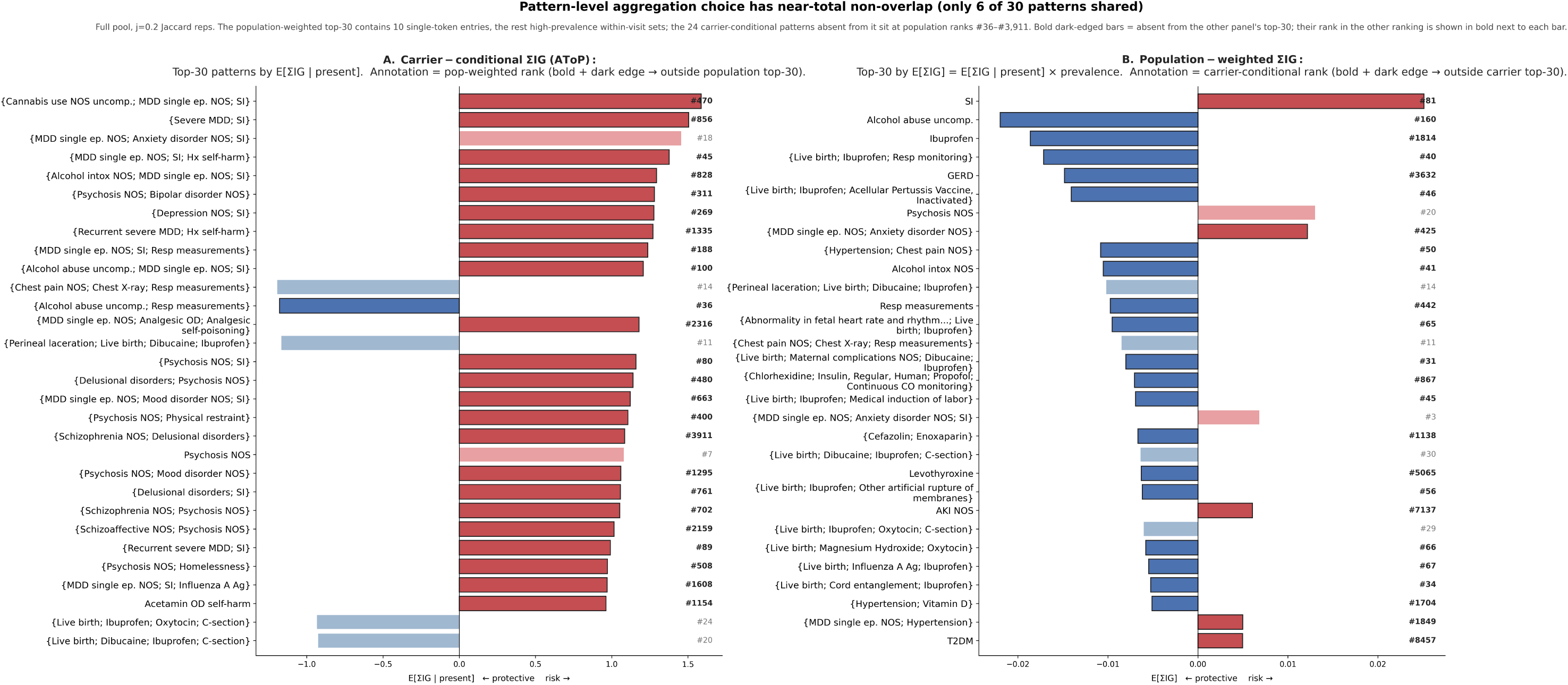
Population-weighted versus carrier-averaged pattern rankings. Top 30 patterns from the Jaccard-deduplicated all-pattern pool (17,891 representatives; threshold 0.20), ranked by (A) carrier-averaged ΣIG and (B) population-weighted attribution. Six patterns appear in both top-30 lists. Dark-outlined bars indicate patterns absent from the other ranking, with their alternative rank shown beside the bar.

**Figure S6.**
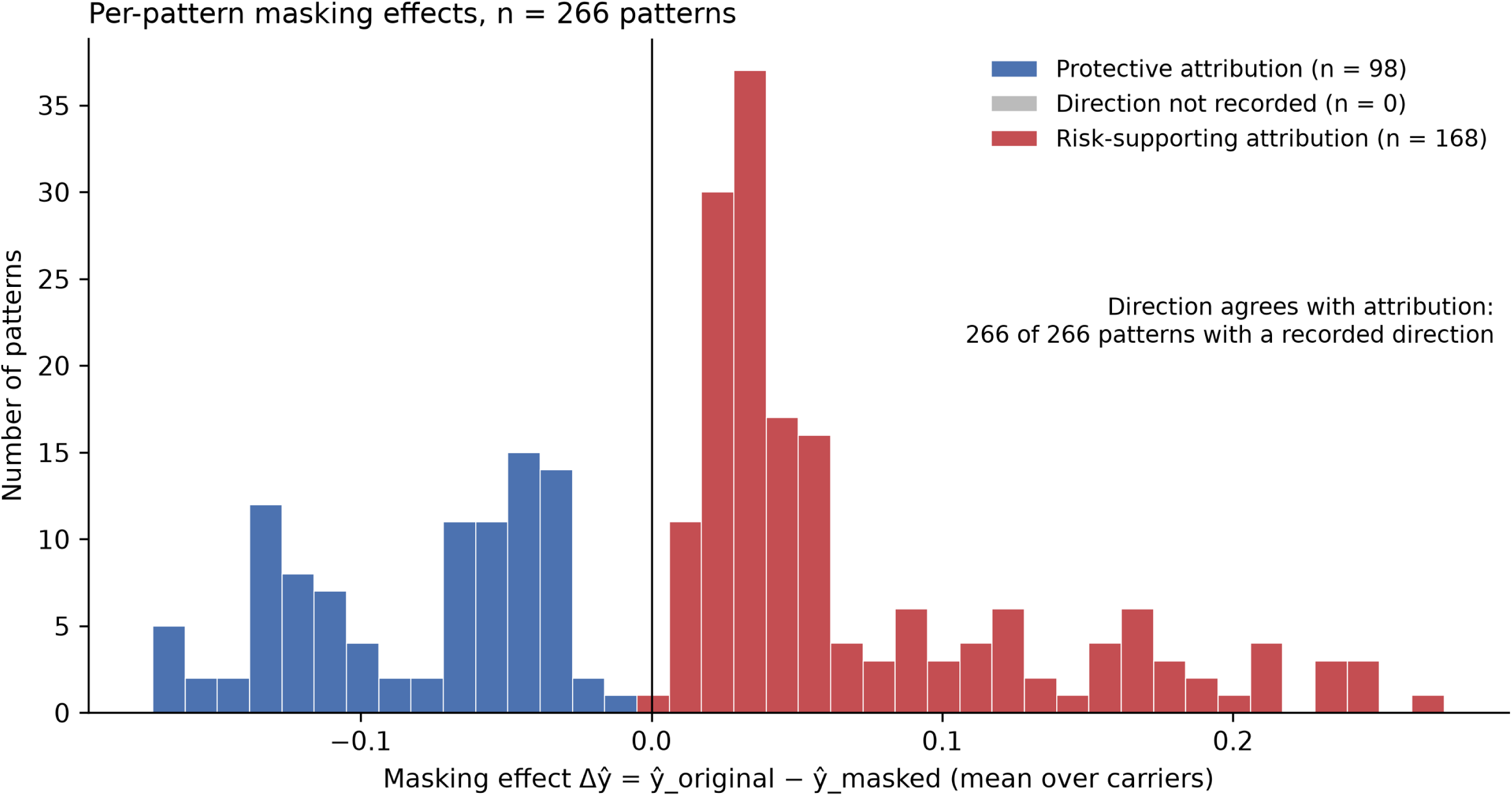
Masking effects across evaluated patterns. Mean prediction change (Δŷ = ŷ_original − ŷ_masked) after masking constituent tokens for 266 patterns, evaluated among up to 200 test-cohort carriers per pattern. Red indicates positive attribution and blue negative attribution based on the original carrier-averaged ΣIG.

**Figure S7.**
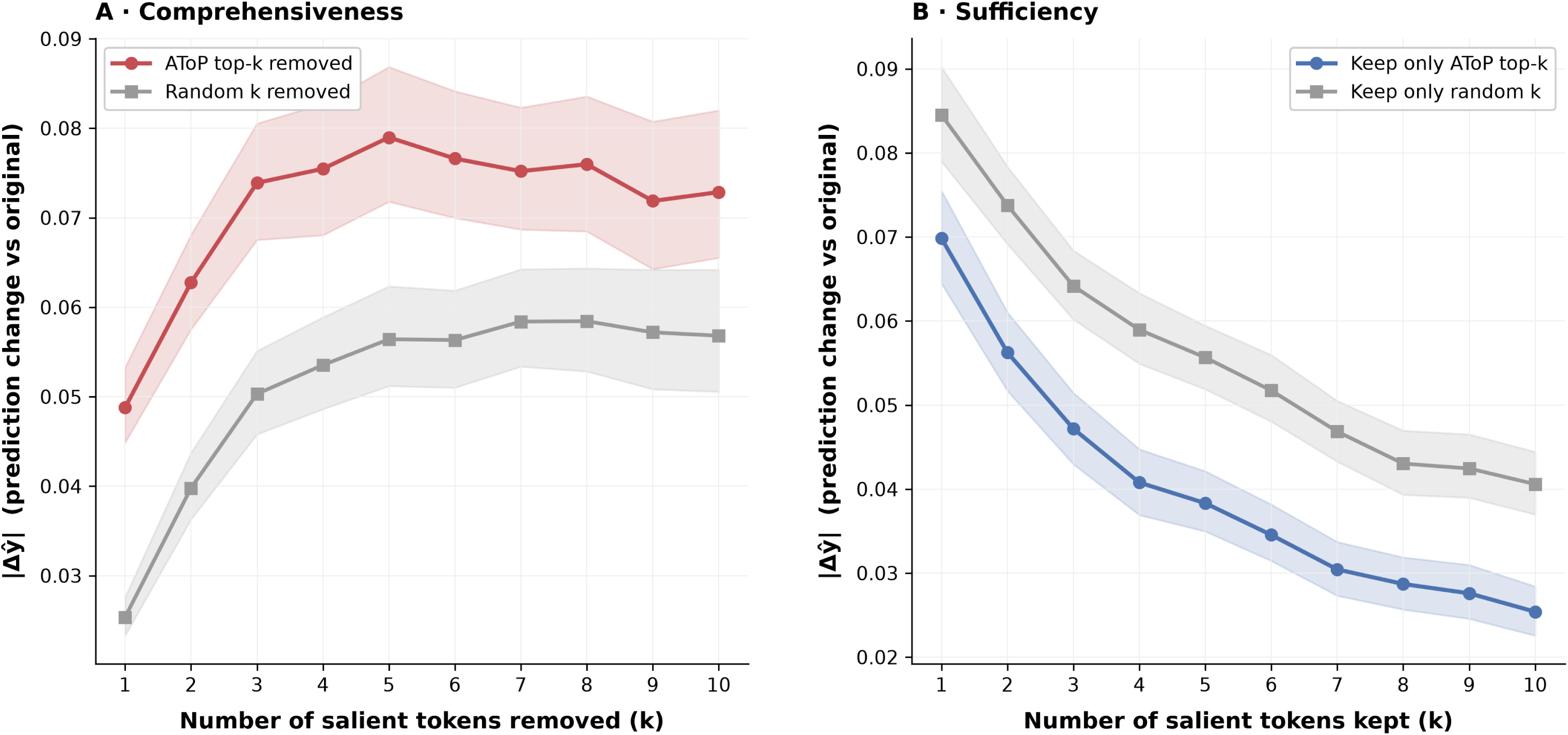
Token-level attribution faithfulness. Changes in predicted readmission probability after selecting the top k tokens by |IG| or an equal number of randomly selected salient tokens (five random draws). Analysis includes 500 sampled test patients with at least three salient tokens; each k includes only patients with at least k. (A) Comprehensiveness: mask the selected tokens. (B) Sufficiency: retain selected tokens and mask the remaining salient tokens. Non-salient and special tokens remain unchanged. Curves show mean absolute prediction change; bands show 95% patient-bootstrap confidence intervals.

**Figure S8.**
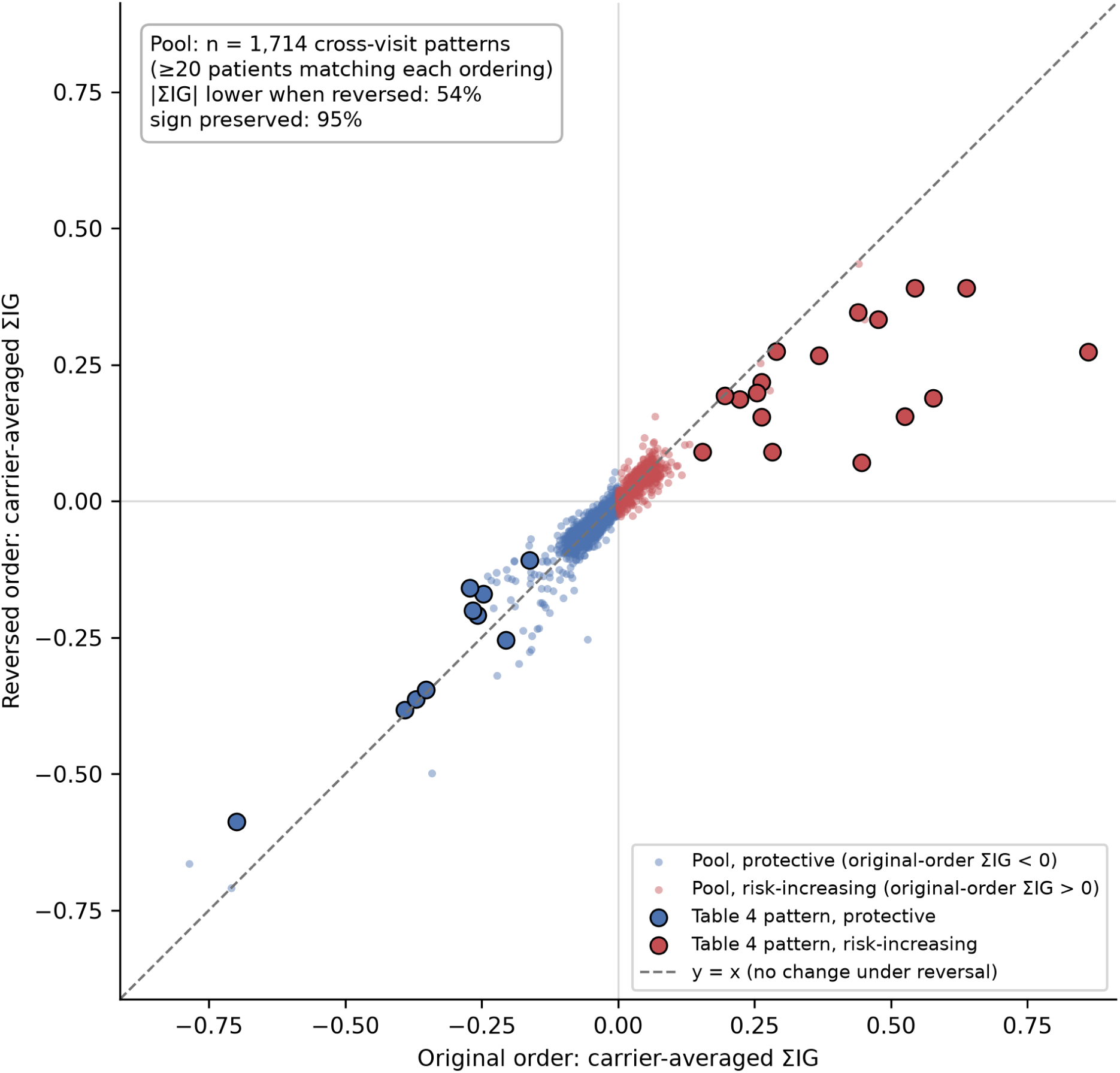
Attribution under original and reversed temporal order. Attribution magnitude decreased for 54% of 1,714 eligible cross-visit patterns in the broader pool, compared with 26 of 27 evaluable patterns (96%) reported in Table 4; attribution sign was preserved for 95% of the broader pool. The horizontal and vertical axes show carrier-averaged signed ΣIG for patients whose attribution-salient visit blocks match the original and reversed orders, respectively. The broader pool required at least 20 matching patients per ordering; outlined points also show the 27 evaluable Table 4 patterns, regardless of this threshold. Three patterns identical under reversal were excluded. Red indicates positive original-order attribution, blue negative attribution, and the dashed diagonal equal attribution. Patient sets may overlap; these between-patient comparisons do not isolate the effect of temporal order.

**Figure S9.**
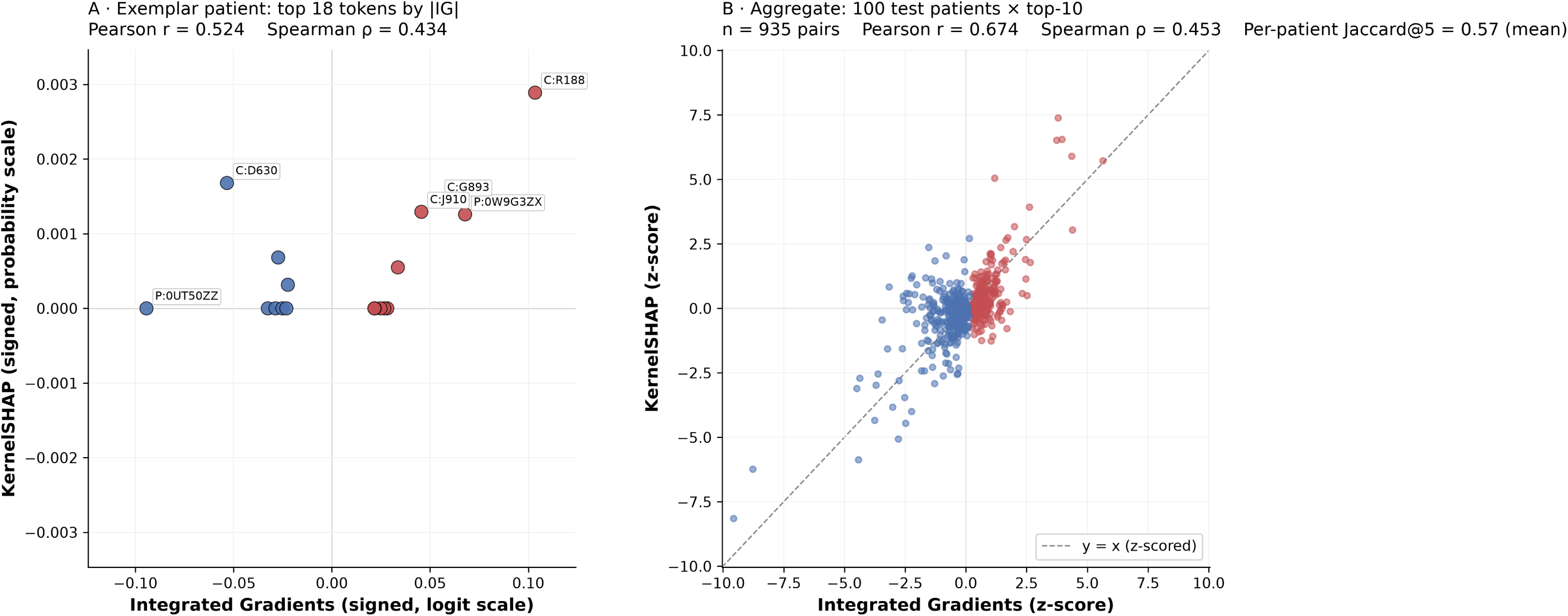
Per-patient agreement between IG and KernelSHAP. (A) Signed token attributions for a psychiatric true-positive patient (18 top-ranked IG tokens; six labeled). (B) Attributions for the 10 highest-|IG| tokens from each of 100 sampled patients (935 patient-token pairs), standardized within method. Red indicates positive IG attribution and blue negative attribution; the dashed line shows equality. IG is calculated on the logit and KernelSHAP on predicted probability. KernelSHAP used 200 coalitions per patient here, compared with approximately 50 for cohort-level analyses in Figures S10–S11. Agreement was Pearson r = 0.52 and Spearman ρ = 0.43 in panel A, and r = 0.67 and ρ = 0.45 in panel B; mean Jaccard@5 (top-five token-set overlap) was 0.57.

**Figure S10.**
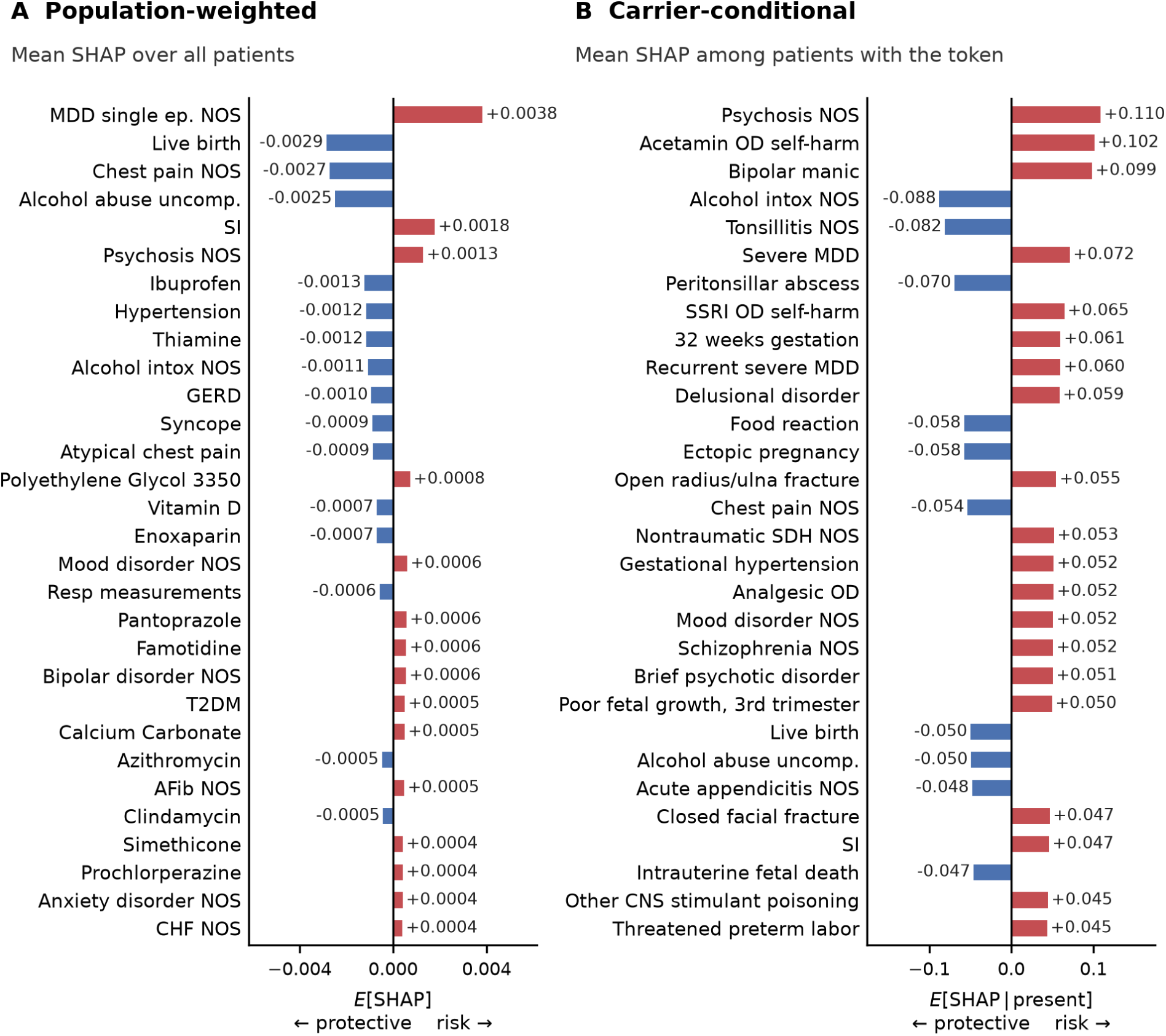
Token rankings using KernelSHAP. Top 30 tokens by absolute mean SHAP attribution across (A) all 43,602 test patients with valid sequences and (B) carriers of each token. Tokens with fewer than 10 sampled carriers were excluded. The carrier-conditional top 30 shared 25 tokens with the IG ranking in Figure 3.

**Figure S11.**
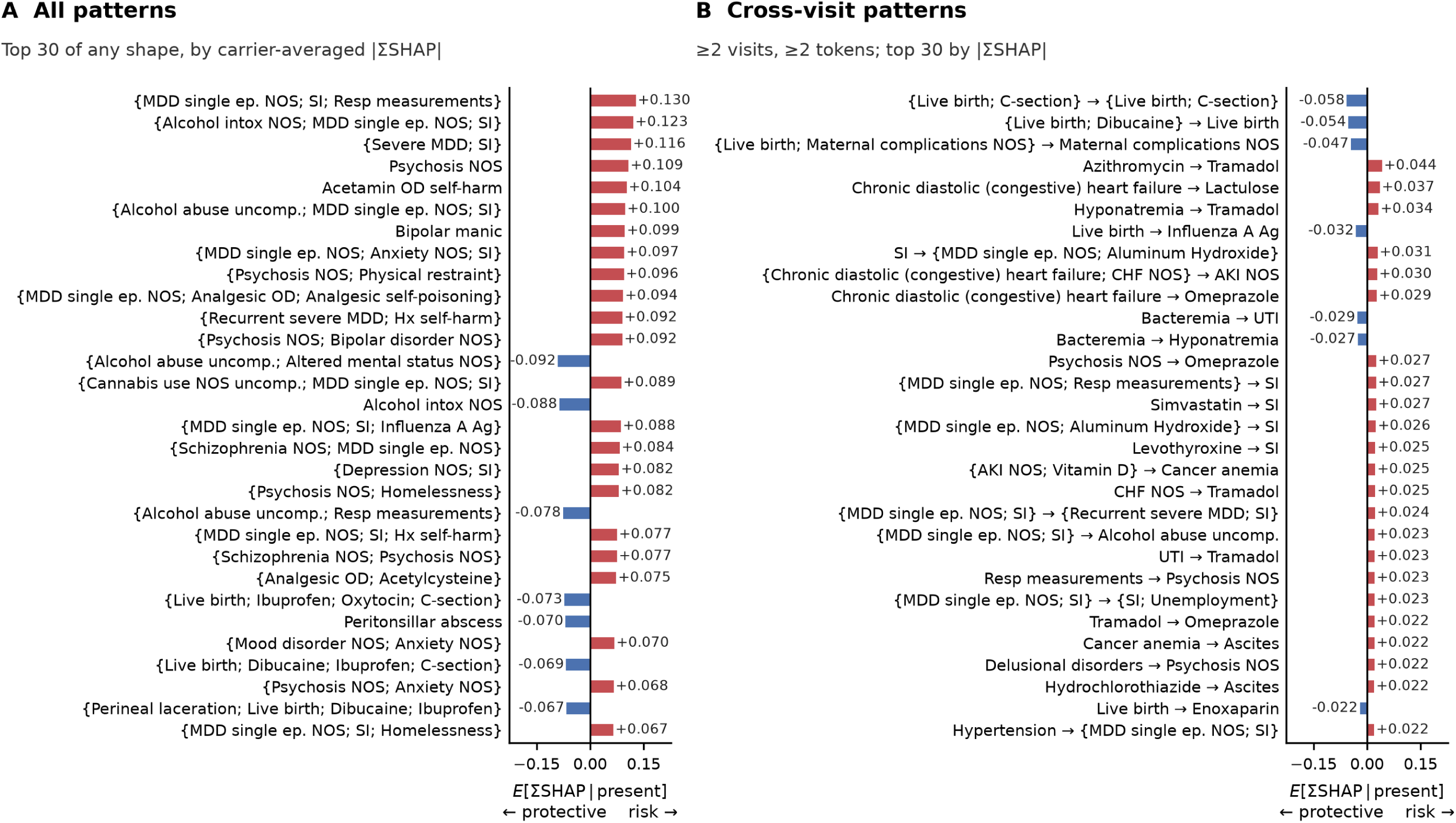
Pattern rankings using KernelSHAP. Top 30 patterns by absolute carrier-averaged ΣSHAP for (A) all pattern types and (B) cross-visit sequences. KernelSHAP scores were applied to the pattern representatives selected using IG-based Jaccard deduplication (threshold 0.20); representatives were not independently reselected by SHAP magnitude. Patterns with fewer than 10 sampled carriers were excluded. The rankings shared 21 of 30 all-pattern and 7 of 30 cross-visit patterns with IG (Figure 4).

**Figure S12.**
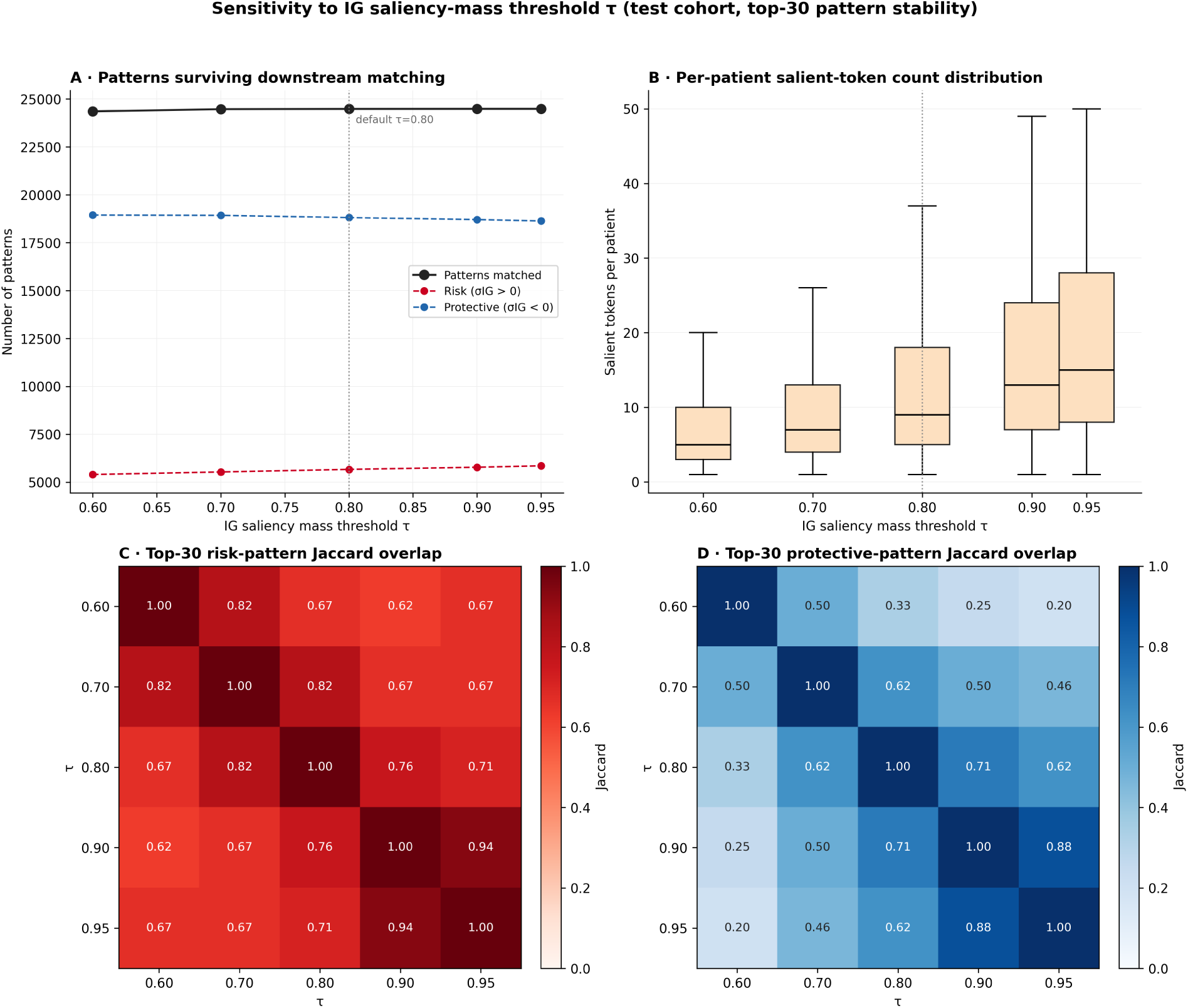
Sensitivity to the IG saliency-mass threshold. The threshold τ was varied from 0.60 to 0.95 (default 0.80) while keeping the mined pattern dictionary fixed and re-matching patterns. (A) Number of patterns matched to at least one test patient. (B) Number of salient tokens retained per patient. (C) Pairwise top-30 Jaccard overlap for positively attributed patterns. (D) Pairwise top-30 Jaccard overlap for negatively attributed patterns.

**Table S1.** Transformer architecture and training settings.

| Section | Parameter | Value |
|---|---|---|
| Input | Vocabulary size | 24,005 tokens (14,960 diagnosis; 7,877 procedure; 1,165 RxNorm ingredient; 3 special) |
|  | Special tokens | [PAD]=0, [CLS]=1, [VISIT]=2 |
|  | Within-visit token order | diagnoses by seq_num, then procedures by seq_num, then first-recorded prescriptions by start time |
|  | Visit window | index admission plus up to 19 preceding admissions (max_visits = 20) |
|  | Maximum sequence length | 512 tokens |
|  | Records truncated | 7 of 218,196 (0.003%): train 4 / 152,737; validation 1 / 21,819; test 2 / 43,640 |
|  | Out-of-vocabulary tokens | mapped to [PAD], excluded by the key-padding mask, position slot retained |
| Architecture | Embedding dimension | 128 |
|  | Positional encoding | learned absolute position embedding (512 x 128) added to the token embedding, then dropout and LayerNorm; no visit-, type- or time-level embeddings |
|  | Encoder layers | 2, pre-LayerNorm (norm_first), no final LayerNorm |
|  | Attention heads | 4 (32 dimensions per head) |
|  | Feed-forward dimension | 512 (4 x 128), ReLU activation |
|  | Dropout | 0.1 (embedding, attention, feed-forward, classifier) |
|  | Classification head | [CLS] output -> Linear 128->64 -> ReLU -> Dropout -> Linear 64->1 -> sigmoid |
|  | Trainable parameters | 3,543,297 (token embeddings 3,072,640; position embeddings 65,536; encoder 396,544; head 8,321; embedding LayerNorm 256) |
| Training | Loss | binary cross-entropy with logits, positive-class weight 6.91 (= 133,420 negatives / 19,317 positives in the training split) |
|  | Optimizer | AdamW, beta = (0.9, 0.999), epsilon = 1e-8 (library defaults) |
|  | Learning rate | 1e-4, constant (no warm-up, no schedule) |
|  | Weight decay | 1e-5 |
|  | Gradient clipping | none |
|  | Batch size | 64 (training data shuffled each epoch) |
|  | Maximum epochs | 50 |
|  | Early stopping | validation AUROC, patience 5, best weights restored |
|  | Epochs run / selected | 13 / 8 (validation AUROC 0.7585, PR-AUC 0.3217 at epoch 8) |
|  | Random seed | 42 (Python, NumPy, PyTorch, CUDA; cuDNN deterministic); single training run, no seed-variability runs |
|  | Data split | patient-level random 70/10/20, seed 42: 152,737 / 21,819 / 43,640 patients; one record per patient; not stratified by outcome (prevalence 12.65% / 12.06% / 12.74%); not temporal |
|  | Hardware and software versions | not recorded in the run bundle |

**Abbreviations:** AUROC = area under the receiver operating characteristic curve; OOV = out of vocabulary; PR-AUC = area under the precision-recall curve. **Note:** Settings absent from the saved configuration reflect training-script defaults. Hardware and library versions were not recorded.

**Table S2.** AToP parameter and evaluation settings.

| **Parameter** | **Value** | **Selection** | **Rationale or sensitivity** |
|---|---|---|---|
| Maximum drug prevalence | 0.25 | Selected by inspection | Removes near-universal inpatient agents; set by inspecting training-cohort patterns without outcome labels |
| Saliency-mass threshold (τ) | 0.80 | Prespecified | Captures 80% of absolute attribution mass; sensitivity across 0.60 to 0.95 (Figure S12) |
| Maximum salient tokens per patient | 50 | Prespecified | Tractability cap; binding for 1,812 of 43,602 test patients (4.2%) |
| Mining minimum support | 0.001 per class | Prespecified | At least 0.1% of training patients per class |
| Maximum pattern length | 4 tokens | Prespecified | Bounds candidate generation |
| Jaccard deduplication threshold (j) | 0.20 | Prespecified | Reduces overlapping patterns; sensitivity in Figures S3 and S4 |
| Cross-visit ranking: minimum visits and tokens | 2 visits; 2 distinct tokens | Prespecified | Restricts the ranking to longitudinal sequences |
| Aggregation convention | Carrier-conditional | Design choice | Population-weighted shown for contrast (Figures 3, S5) |
| Bootstrap resamples for performance CIs | 1,000 | Prespecified | Percentile intervals, seed 42 (Table 2) |
| IG integration steps | 30 | Prespecified | Median completeness residual 1.8% (Supplementary Methods) |
| Per-patient explanation dictionary | Support ≥0.001 of training cohort | Prespecified | 24,490 of 70,278 pooled episodes (Table S3) |
| Reporting floor, all-patterns ranking | ≥5 test-cohort carriers | Prespecified | Implementation default; not tuned |
| Reporting floor, cross-visit ranking | ≥3 test-cohort carriers | Prespecified | Implementation default; not tuned |
| Masking validation: carriers per pattern | ≤200 | Prespecified | 266 unique patterns (Figure S6) |
| Within-patient reversal: carriers per pattern | ≤80 | Prespecified | Table S6 |
| Faithfulness: patients, random draws, bootstrap resamples | 500; 5; 1,000 | Prespecified | Patients with 3 or more salient tokens; seed 42 (Figure S7) |
| KernelSHAP coalitions per patient | ≈50 (200 for Figure S9) | Prespecified | Tractability on 43,602 patients |

**Abbreviations:** CI = confidence interval; IG = Integrated Gradients; j = Jaccard similarity threshold; τ = saliency-mass threshold. **Note:** “Prespecified” means fixed before the corresponding analysis; “design choice” denotes a methodological convention. The drug-prevalence threshold alone was selected by inspecting training-cohort output, without test outcomes.

**Table S3.** AToP pattern mining pipeline summary.

| **Statistic** | **Value** |
|---|---|
| Attribution (training cohort) |  |
| Sequence length, tokens, mean (SD) | 33.2 (41.3) |
| Salient tokens, 80% IG mass, mean (SD) | 14.8 (16.7) |
| Token compression ratio (full / salient) | 2.2 |
| Frequent-episode mining (training cohort, support 0.001 within each outcome class) |  |
| Episodes mined in readmitted patients (≥20 carriers) | 67,321 |
| Episodes mined in non-readmitted patients (≥134 carriers) | 21,650 |
| Distinct episodes after pooling | 70,278 |
| Pooled episodes also at whole-cohort support 0.001 (≥153 carriers; per-patient explanation dictionary) | 24,490 |
| Composition of the per-patient dictionary |  |
| Single-token patterns | 1,602 |
| Within-visit co-occurrence patterns | 17,563 |
| Cross-visit patterns | 5,325 |
| Candidate patterns scored on test cohort (the 70,278 pooled episodes) |  |
| Full pool, all shapes, ≥5 test carriers | 70,275 |
| Cross-visit candidates (≥2 visits, ≥2 distinct tokens; ≥3 test carriers) | 39,611 |
| Representatives after Jaccard dedup (j = 0.20, test carriers) |  |
| Full pool (all shapes) | 17,891 |
| Cross-visit (locked reporting dictionary) | 9,213 |
| Risk, ΣIG > 0 | 3,741 |
| Protective, ΣIG < 0 | 5,472 |

**Abbreviations:** IG = Integrated Gradients; SD = standard deviation; ΣIG = sum of token IG attributions. **Note:** Patterns passing the support threshold in either training outcome class were pooled for global ranking; the per-patient dictionary additionally required support in the full training cohort. Counts represent distinct pipeline stages, not a running total. Token compression ratio is mean full-sequence length divided by mean salient-subsequence length. Test carriers were matched against complete records.

**Table S4.** Association of cross-visit patterns with readmission, adjusting for the number of admissions.

| **Clinical category** | **Pattern (representative)** | **ΣIG** | **n** | **Carrier readmit %** | **Non-carrier readmit %** | **Adj. OR** | **95% CI** |
|---|---|---|---|---|---|---|---|
| Psychiatric | {MDD single ep. NOS; SI} → {Recurrent severe MDD; SI} | +0.80 | 36 | 44.4% | 27.7% | 1.91 | 0.99–3.71 |
|  | {MDD single ep. NOS; SI} → {SI; Unemployment NOS} | +0.60 | 10 | 30.0% | 27.8% | 0.94 | 0.24–3.64 |
|  | {MDD single ep. NOS; SI} → {Anxiety disorder NOS; SI} | +0.59 | 37 | 51.4% | 27.7% | 2.43 | 1.27–4.65 |
|  | {MDD single ep. NOS; SI} → SI | +0.55 | 201 | 42.3% | 27.5% | 1.71 | 1.28–2.27 |
|  | {MDD single ep. NOS; SI} → {SI; Thiamine} | +0.53 | 29 | 34.5% | 27.8% | 1.22 | 0.57–2.64 |
| Oncologic | Bone metastases → Bone metastases → {Cancer anemia; Bone metastases} | +0.55 | 13 | 69.2% | 27.7% | 5.56 | 1.71–18.09 |
|  | Cancer anemia → Ascites | +0.48 | 21 | 76.2% | 27.7% | 7.44 | 2.72–20.36 |
|  | Cancer anemia → Cancer anemia → {Cancer anemia; Cancer pain} | +0.44 | 21 | 57.1% | 27.7% | 2.84 | 1.19–6.76 |
|  | {Cancer pain; Bone metastases} → {Cancer pain; Bone metastases} | +0.41 | 16 | 56.2% | 27.8% | 3.24 | 1.21–8.73 |
|  | Malignant pleural effusion → Cancer anemia | +0.39 | 14 | 71.4% | 27.7% | 6.29 | 1.96–20.12 |
| Obstetric | {Live birth; C-section} → {Live birth; C-section} | -0.79 | 51 | 23.5% | 27.8% | 0.83 | 0.44–1.60 |
|  | {Live birth; Dibucaine} → Live birth | -0.76 | 165 | 11.5% | 28.0% | 0.35 | 0.22–0.57 |
|  | {Live birth; Maternal complications NOS} → Maternal complications NOS | -0.35 | 23 | 17.4% | 27.8% | 0.54 | 0.18–1.59 |
|  | Live birth → Influenza A Ag | -0.28 | 105 | 19.0% | 27.9% | 0.61 | 0.38–1.00 |
|  | Oxazepam → Maternal comorbidity | +0.25 | 25 | 32.0% | 27.8% | 1.15 | 0.50–2.68 |
| Chest pain workup | {Chest pain NOS; Resp measurements} → Chest pain NOS | -0.37 | 46 | 17.4% | 27.8% | 0.47 | 0.22–1.00 |
|  | Chest pain NOS → Simvastatin | -0.27 | 73 | 21.9% | 27.8% | 0.67 | 0.38–1.16 |
|  | Chest pain NOS → Chest pain NOS → Atypical chest pain | -0.26 | 35 | 17.1% | 27.8% | 0.41 | 0.17–1.00 |
|  | Chest pain NOS → Clindamycin | -0.25 | 39 | 33.3% | 27.8% | 1.04 | 0.53–2.04 |
|  | Chest pain NOS → Chest pain NOS → GERD | -0.23 | 32 | 28.1% | 27.8% | 0.79 | 0.37–1.73 |
| Hepatobiliary | {Obstruction of bile duct; ERCP biliary stent} → Obstruction of bile duct | +0.27 | 57 | 36.8% | 27.8% | 1.38 | 0.80–2.38 |
|  | Obstruction of bile duct → Percutaneous biliary drainage | +0.25 | 17 | 58.8% | 27.7% | 3.28 | 1.24–8.68 |
|  | {GERD; Obstruction of bile duct} → Obstruction of bile duct | +0.16 | 28 | 35.7% | 27.8% | 1.36 | 0.63–2.96 |
|  | Acute pancreatitis NOS → Other chronic pain → Other chronic pain | -0.15 | 24 | 20.8% | 27.8% | 0.51 | 0.19–1.38 |
|  | Obstruction of bile duct → Intrahepatic bile duct carcinoma | +0.15 | 22 | 59.1% | 27.7% | 3.43 | 1.46–8.06 |
| Other | CVC placement → Venous catheterization, NEC → CVC placement → CVC placement | +0.24 | 19 | 31.6% | 27.8% | 0.99 | 0.38–2.62 |
|  | CVC placement → {Thiamine; CVC placement} → CVC placement | +0.23 | 7 | 14.3% | 27.8% | 0.37 | 0.04–3.05 |
|  | {Thiamine; Vitamin D} → Clindamycin | -0.22 | 27 | 22.2% | 27.8% | 0.64 | 0.26–1.59 |
|  | {Meropenem; CVC placement} → CVC placement → CVC placement | +0.22 | 13 | 38.5% | 27.8% | 1.39 | 0.45–4.25 |
|  | Lactulose → CVC placement → CVC placement → CVC placement | +0.21 | 24 | 29.2% | 27.8% | 0.88 | 0.36–2.13 |

**Abbreviations:** ΣIG = mean signed sum of token attributions among carriers (logit scale); n = pattern carriers in the multi-visit population; Carrier readmit % and Non-carrier readmit % = observed readmission rates in the respective groups; Adj. OR = adjusted odds ratio; CI = confidence interval. **Note:** Each pattern was analyzed among patients with at least two admissions using logistic regression adjusted for log-transformed admission count. Confidence intervals are 95% Wald intervals; no multiplicity correction was applied.

**Table S5.** Original- and reversed-order attribution for the 30 cross-visit patterns in Table 4.

| **Clinical category** | **Pattern (representative)** | **n original** | **n reversed** | **n both** | **ΣIG original** | **ΣIG reversed** | **Attenuation, %** |
|---|---|---|---|---|---|---|---|
| Psychiatric | {MDD single ep. NOS; SI} → {Recurrent severe MDD; SI} | 29 | 14 | 11 | +0.861 | +0.274 | 68.2% |
|  | {MDD single ep. NOS; SI} → SI | 148 | 107 | 83 | +0.638 | +0.390 | 38.8% |
|  | {MDD single ep. NOS; SI} → {SI; Unemployment NOS} | 8 | 9 | 4 | +0.577 | +0.189 | 67.3% |
|  | {MDD single ep. NOS; SI} → {Anxiety disorder NOS; SI} | 10 | 8 | 4 | +0.525 | +0.156 | 70.4% |
|  | {MDD single ep. NOS; SI} → {SI; Thiamine} | 13 | 4 | 2 | +0.446 | +0.071 | 84.1% |
| Oncologic | Bone metastases → Bone metastases → {Cancer anemia; Bone metastases} | 13 | 11 | 4 | +0.544 | +0.391 | 28.2% |
|  | Cancer anemia → Ascites | 21 | 14 | 4 | +0.476 | +0.333 | 30.0% |
|  | Cancer anemia → Cancer anemia → {Cancer anemia; Cancer pain} | 21 | 11 | 8 | +0.439 | +0.346 | 21.2% |
|  | {Cancer pain; Bone metastases} → {Cancer pain; Bone metastases} | 16 | 16 | 16 | +0.405 | +0.405 | 0.0% |
|  | Malignant pleural effusion → Cancer anemia | 13 | 13 | 3 | +0.368 | +0.267 | 27.3% |
| Obstetric | {Live birth; C-section} → {Live birth; C-section} | 44 | 44 | 44 | -0.709 | -0.709 | 0.0% |
|  | {Live birth; Dibucaine} → Live birth | 72 | 2 | 0 | -0.699 | -0.587 | 16.0% |
|  | {Live birth; Maternal complications NOS} → Maternal complications NOS | 19 | 19 | 19 | -0.391 | -0.382 | 2.3% |
|  | Live birth → Influenza A Ag | 49 | 20 | 1 | -0.352 | -0.345 | 2.1% |
|  | Oxazepam → Maternal comorbidity | 19 | 10 | 5 | +0.263 | +0.154 | 41.3% |
| Chest pain workup | {Chest pain NOS; Resp measurements} → Chest pain NOS | 36 | 16 | 14 | -0.370 | -0.363 | 2.0% |
|  | Chest pain NOS → Simvastatin | 45 | 99 | 10 | -0.272 | -0.159 | 41.5% |
|  | Chest pain NOS → Chest pain NOS → GERD | 16 | 30 | 2 | -0.267 | -0.200 | 25.0% |
|  | Chest pain NOS → Clindamycin | 38 | 42 | 5 | -0.257 | -0.209 | 18.9% |
|  | Chest pain NOS → Chest pain NOS → Atypical chest pain | 34 | 44 | 13 | -0.247 | -0.170 | 31.2% |
| Hepatobiliary | {Obstruction of bile duct; ERCP biliary stent} → Obstruction of bile duct | 51 | 20 | 16 | +0.290 | +0.275 | 5.4% |
|  | Obstruction of bile duct → Percutaneous biliary drainage | 15 | 10 | 7 | +0.282 | +0.090 | 68.0% |
|  | {GERD; Obstruction of bile duct} → Obstruction of bile duct | 17 | 7 | 3 | +0.195 | +0.193 | 1.1% |
|  | Acute pancreatitis NOS → Other chronic pain → Other chronic pain | 22 | 12 | 11 | -0.162 | -0.108 | 33.5% |
|  | Obstruction of bile duct → Intrahepatic bile duct carcinoma | 19 | 17 | 10 | +0.155 | +0.090 | 41.9% |
| Other | CVC placement → {Thiamine; CVC placement} → CVC placement | 5 | 5 | 5 | +0.269 | +0.269 | 0.0% |
|  | {Meropenem; CVC placement} → CVC placement → CVC placement | 7 | 4 | 1 | +0.263 | +0.219 | 16.7% |
|  | CVC placement → Venous catheterization, NEC → CVC placement → CVC placement | 11 | 11 | 8 | +0.254 | +0.199 | 21.8% |
|  | Lactulose → CVC placement → CVC placement → CVC placement | 11 | 4 | 0 | +0.222 | +0.187 | 15.9% |
|  | {Thiamine; Vitamin D} → Clindamycin | 14 | 7 | 0 | -0.206 | -0.254 | -23.8% |

**Abbreviations:** n original, n reversed = carriers matching each order; n both = carriers matching both; ΣIG original, ΣIG reversed = mean signed attribution for each order; Attenuation, % = 100 × (1 − \|ΣIG reversed\| / \|ΣIG original\|). **Note:** Order refers to the sequence reported in Table 4. Counts and attribution scores use attribution-salient visit blocks, unlike the full-record results in Table 4. Carrier groups may overlap. Three patterns are unchanged by reversal and were excluded from attenuation summaries. Some reversed-order groups contain very few patients, so their attenuation estimates are descriptive.

**Table S6.** Within-patient temporal-order reversal for the 30 cross-visit patterns in Table 4.

| Clinical category | Pattern | n | ΣIG original | ΣIG reversed | Attenuation, % | Patients with reduced \|ΣIG\|, % | Attribution sign preserved, % | P value |
|---|---|---|---|---|---|---|---|---|
| Psychiatric | {MDD single ep. NOS; SI} → {Recurrent severe MDD; SI} | 36 | +0.795 | +0.785 | 1.3 | 50.0 | 100 | 0.515 |
|  | {MDD single ep. NOS; SI} → SI | 80 | +0.549 | +0.385 | 29.9 | 73.8 | 92 | <0.001 |
|  | {MDD single ep. NOS; SI} → {SI; Unemployment NOS} | 10 | +0.604 | +0.492 | 18.5 | 40.0 | 100 | 0.461 |
|  | {MDD single ep. NOS; SI} → {Anxiety disorder NOS; SI} | 37 | +0.595 | +0.441 | 25.9 | 67.6 | 89 | 0.006 |
|  | {MDD single ep. NOS; SI} → {SI; Thiamine} | 29 | +0.529 | +0.424 | 19.8 | 62.1 | 86 | 0.055 |
| Oncologic | Bone metastases → Bone metastases → {Cancer anemia; Bone metastases} | 13 | +0.549 | +0.428 | 22.2 | 76.9 | 100 | 0.034 |
|  | Cancer anemia → Ascites | 21 | +0.476 | +0.465 | 2.4 | 57.1 | 100 | 0.317 |
|  | Cancer anemia → Cancer anemia → {Cancer anemia; Cancer pain} | 21 | +0.439 | +0.360 | 18.1 | 71.4 | 100 | 0.084 |
|  | {Cancer pain; Bone metastases} → {Cancer pain; Bone metastases} | 16 | +0.406 | +0.539 | -32.7 | 31.2 | 100 | 0.986 |
|  | Malignant pleural effusion → Cancer anemia | 14 | +0.392 | +0.367 | 6.3 | 50.0 | 100 | 0.357 |
| Obstetric | {Live birth; C-section} → {Live birth; C-section} | 51 | -0.791 | -0.726 | 8.2 | 56.9 | 100 | 0.077 |
|  | {Live birth; Dibucaine} → Live birth | 80 | -0.759 | -0.722 | 4.8 | 55.0 | 100 | 0.263 |
|  | {Live birth; Maternal complications NOS} → Maternal complications NOS | 23 | -0.351 | -0.288 | 17.9 | 52.2 | 100 | 0.136 |
|  | Live birth → Influenza A Ag | 80 | -0.302 | -0.350 | -15.9 | 40.0 | 100 | 0.983 |
|  | Oxazepam → Maternal comorbidity | 25 | +0.252 | +0.265 | -5.1 | 56.0 | 100 | 0.500 |
| Chest pain workup | {Chest pain NOS; Resp measurements} → Chest pain NOS | 46 | -0.370 | -0.340 | 8.2 | 60.9 | 98 | 0.041 |
|  | Chest pain NOS → Simvastatin | 73 | -0.269 | -0.143 | 46.8 | 74.0 | 94 | <0.001 |
|  | Chest pain NOS → Chest pain NOS → GERD | 32 | -0.232 | -0.151 | 35.1 | 81.2 | 91 | <0.001 |
|  | Chest pain NOS → Clindamycin | 39 | -0.251 | -0.201 | 20.0 | 64.1 | 100 | 0.022 |
|  | Chest pain NOS → Chest pain NOS → Atypical chest pain | 35 | -0.256 | -0.229 | 10.7 | 65.7 | 100 | 0.113 |
| Hepatobiliary | {Obstruction of bile duct; ERCP biliary stent} → Obstruction of bile duct | 57 | +0.270 | +0.298 | -10.5 | 43.9 | 100 | 0.904 |
|  | Obstruction of bile duct → Percutaneous biliary drainage | 17 | +0.254 | +0.070 | 72.5 | 94.1 | 94 | <0.001 |
|  | {GERD; Obstruction of bile duct} → Obstruction of bile duct | 28 | +0.158 | +0.174 | -10.4 | 50.0 | 93 | 0.819 |
|  | Acute pancreatitis NOS → Other chronic pain → Other chronic pain | 24 | -0.152 | -0.092 | 39.6 | 66.7 | 100 | 0.010 |
|  | Obstruction of bile duct → Intrahepatic bile duct carcinoma | 22 | +0.146 | +0.136 | 7.1 | 63.6 | 100 | 0.138 |
| Other | CVC placement → {Thiamine; CVC placement} → CVC placement | 7 | +0.228 | +0.173 | 24.1 | 71.4 | 100 | 0.039 |
|  | {Meropenem; CVC placement} → CVC placement → CVC placement | 13 | +0.217 | +0.240 | -10.8 | 46.2 | 100 | 0.729 |
|  | CVC placement → Venous catheterization, NEC → CVC placement → CVC placement | 19 | +0.236 | +0.195 | 17.2 | 73.7 | 100 | 0.067 |
|  | Lactulose → CVC placement → CVC placement → CVC placement | 24 | +0.206 | +0.226 | -9.8 | 45.8 | 100 | 0.658 |
|  | {Thiamine; Vitamin D} → Clindamycin | 27 | -0.217 | -0.167 | 22.9 | 51.9 | 100 | 0.235 |

**Abbreviations:** n = carriers tested (up to the first 80 per pattern); ΣIG original, ΣIG reversed = mean attribution before and after reordering matched visits and recomputing IG; Attenuation, % = 100 × (1 − |ΣIG reversed| / |ΣIG original|); Patients with reduced |ΣIG|, % = carriers with reduced attribution magnitude; Attribution sign preserved, % = carriers retaining the sign; P value = one-sided Wilcoxon signed-rank test over carriers. **Note:** Unlike Table S5, this test reorders matched whole visits within each patient's record. Other events in those visits move with them, so the analysis does not isolate the order of the pattern tokens. Three patterns are symmetric under reversal; changes in their scores reflect other visit contents, and they were excluded from aggregate summaries. Per-pattern P values are nominal; 4 of 27 remained significant after Holm correction.

## Notes

### Competing Interest Statement

The authors have declared no competing interest.

### Author Declarations

This study used the de-identified MIMIC-IV v3.1 database (Beth Israel Deaconess Medical Center, Boston, MA, USA), accessed under the PhysioNet credentialed data use agreement after completion of the required CITI training. The creation and sharing of MIMIC-IV was approved by the Institutional Review Board of Beth Israel Deaconess Medical Center, which granted a waiver of informed consent. Secondary analysis of this de-identified resource did not require separate institutional review board approval.

